# Unmasking Hidden Dysglycemia: A Mobile OGTT Approach Using Continuous Glucose Monitors

**DOI:** 10.64898/2025.12.14.25341915

**Authors:** Karnika Singh, Lucy Esteve, Penelope North, Anna Beason, Leeor Hershkovich, Bill Chen, Shekh Md Mahmudul Islam, Brinnae Bent, Peter J. Cho, Will Ke Wang, Md Mobashir Hasan Shandhi, Michael P. Snyder, Ahmed A. Metwally, Matthew J. Crowley, Anastasia-Stefania Alexopoulos, Jessilyn Dunn

## Abstract

Prediabetes (PD), which includes impaired glucose tolerance (IGT) and impaired fasting glucose (IFG), is a dysfunctional metabolic state that often progresses to type 2 diabetes (T2D). Standard screening tools such as random glucose and hemoglobin A1c frequently miss early or intermittent dysglycemia and cannot distinguish underlying physiological differences relevant for targeted intervention. Although the oral glucose tolerance test (OGTT) detects more PD and T2D and identifies high-risk individuals earlier, its clinical use is limited by the need for repeated venous sampling and in-clinic administration. We introduce the mobile OGTT (mOGTT), which leverages continuous glucose monitoring (CGM) to capture high-resolution glycemic responses to a standardized glucose challenge outside clinical settings. In a population spanning a broad range of glycemic health, we establish preliminary normative mOGTT values, characterize their relationship to clinical OGTT thresholds, and assess concordance with A1c. We show that CGM-derived analogs of OGTT metrics improve detection and phenotyping of dysglycemia and propose the Glucose Challenge Response Index (GCRI), a composite measure of glycemic health. Finally, we demonstrate the generalizability of GCRI-based subphenotypes in an out-of-sample cohort. These results help facilitate an efficient and scalable approach for conveniently detecting and quantifying early-stage glucose dysregulation.

## Introduction

Metabolic impairments such as impaired insulin secretion (i.e., impaired glucose tolerance; IGT), impaired fasting glucose (IFG), and insulin resistance are early hallmarks of prediabetes (PD) and type 2 diabetes (T2D) but are easily missed through hemoglobin A1c (or A1c) testing. Because β-cell function is already substantially compromised once A1c thresholds for PD or T2D are crossed, A1c testing misses early IFG and IGT and also fails to identify differential underlying pathophysiologies of glycemic dysfunction for personalized intervention.^2^

The oral glucose tolerance test (OGTT) enables clinical assessment of IGT and involves monitoring a person’s glucose response to a consumed glucose load (usually 75 grams) and measuring blood glucose at predetermined intervals (typically 1- or 2-hours post-ingestion). Typical OGTT diagnostic methods compare fasting plasma glucose (FPG) and interval glucose measurements against normative values.^1^ Of these measurements, the 30-min and 1-hour OGTT glucose levels are increasingly being recognized for their value in detecting earlier signs of dysglycemia as compared with FPG, 2-hour OGTT glucose levels, or A1c.^3^ Such early measures can capture signs of risk in individuals who still have their β-cell function largely intact, opening a critical window of opportunity for preventive intervention.^3^ Because OGTT has greater sensitivity for detecting T2D and PD than A1c ^4–8^, the International Diabetes Federation now recommends OGTT over A1c and FPG for dysglycemia screening because it can identify high-risk patients earlier.^8^ However, the most appropriate glucose measurement time points and diagnostic threshold values remain under discussion.^3^

Despite OGTT being recognized as the most sensitive test to detect IGT, PD, and T2D ^7–9^, it is rarely used by clinicians or patients because its procedures are burdensome, i.e., it requires multiple serum glucose measurements, with patients having to wait at the clinic/hospital setting for the duration of the study, as per standard practice. Thus, while OGTT provides critical information about a person’s glycemic health that cannot be captured through common clinical approaches, it is not routinely used as a method to screen for early metabolic impairments.

The OGTT glucose challenge response is shaped by the rate of carbohydrate absorption, the secretion of insulin and glucagon, and their coordinated regulation of hepatic and peripheral glucose metabolism.^10^ OGTT dynamics (e.g., monophasic, biphasic, or continuously rising), while not accessible through traditional sampling methods or used in routine practice, has the potential to identify root causes of dysglycemia, for example, insulin resistance or β-cell dysfunction.^11^ For example, monophasic and continuous-rise curves are associated with lower insulin sensitivity/secretion and decreased β-cell function with a higher risk of progression to T2D, even when fasting and 2-h glucose remain sub-diagnostic.^11–16^ Novel metrics to characterize OGTT dynamics have recently been explored. For example, multiple point measurements over time can be integrated to gain insight into the total post-load glucose response, quantified as the area under the curve (AUC)^17^, and time to plasma glucose peak has been found to be a useful metric to enhance PD risk stratification.^18^ However, implementation of such metrics has been limited to research settings given the impracticality of serial blood glucose measurements in the clinic.

Continuous glucose monitors (CGMs) provide an opportunity to obtain higher frequency glucose measurements. These can be used to generate novel metrics of dysglycemia from the glucose response curve during an OGTT, for instance a composite metric like the GCRI introduced here, which combines multiple OGTT metrics and their respective clinical thresholds. However, no consensus method has been developed using CGM metrics derived from OGTT. Further, CGMs confer the added benefits of allowing for glycemic metrics to be assessed outside clinical settings, and also enable repeated testing over time.^19–23^ Despite their promise, CGMs are yet to be routinely adopted for OGTT. A major limitation to realizing CGM-based OGTTs for clinical decision-making is the lack of characteristic CGM data across the glycemic spectrum, which would be essential for establishing reference values and clinically meaningful thresholds to support the translation of CGM-based OGTT into practice. As CGMs gain traction among individuals without diagnosed diabetes, CGM-based (i.e., at-home or “mobile”) OGTTs may offer a clinically grounded, scalable method for longitudinal self-monitoring and early risk detection.

Overall, current reliance on infrequent A1c testing routinely misses changes in glycemic health, such as progression to PD, or even early stages of T2D. While CGMs allow continuous at-home monitoring, no standardized framework exists to assess glycemic health, particularly glucose clearance dynamics. Existing passive CGM metrics are confounded by behavior and context, and no study has defined OGTT-derived values across people with and without PD and T2D. Our work is a starting point for establishing normative mOGTT values by proposing data-driven thresholds for classification (Figure 1 A, B). We further introduce the GCRI, a composite score of glycemic health, laying the foundation for CGM-based home evaluation, progression monitoring, and early detection of PD and T2D (Figure 1C, D).

**Figure 1.**
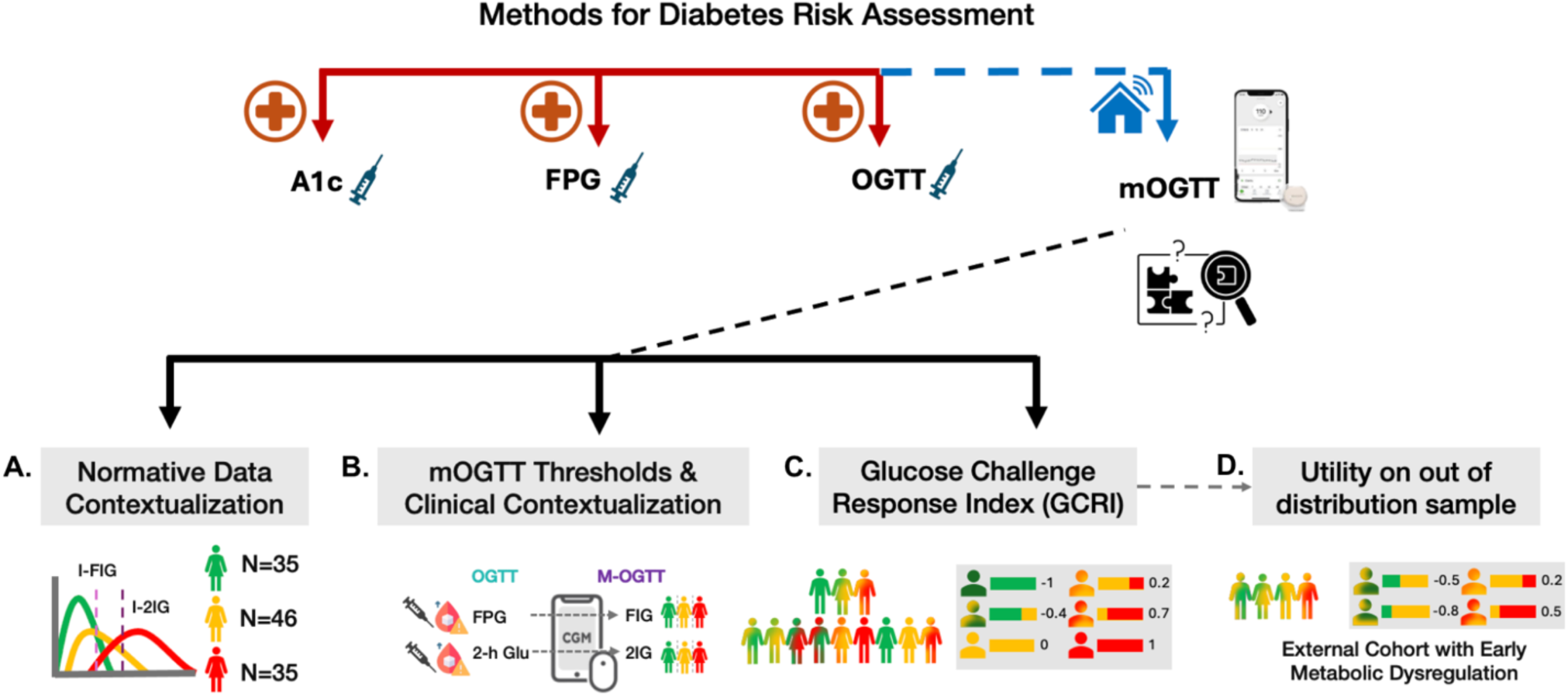
Graphical abstract. Current clinical practice relies on A1c, FPG, or OGTT measurements that require a person’s presence in a clinical setting. Our work is focused on developing a standardized framework to assess glycemic health outside of the clinic by leveraging newly widely available CGMs. Our approach involved ***(A)*** defining normative mOGTT values across Norm, PD, and untreated T2D, ***(B)*** establishing data-driven thresholds for classification, ***(C)*** introducing the GCRI as a composite index of glycemic health, and ***(D)*** validating the GCRI on an external population. Together, these advances support CGM-based mOGTT for home monitoring, progression tracking, and early detection of glycemic dysfunction.

## Results

### DiabetesWatch Cohort Characteristics

Continuous glucose monitor (Abbott Libre 3 CGM) and smartwatch data were collected for two weeks from 116 individuals (N=62 female, N=54 male; N=23 Hispanic/Latino, N=30 Black) participating in the DiabetesWatch study (Duke IRB Pro00112384). Of the 116 participants, 35 had type 2 diabetes (T2D) as defined by an A1c of ≥ 6.5% at baseline, 46 had prediabetes (PD) (A1c 5.7-6.4%), and 35 had normoglycemia (A1c <5.7%). A1c values ranged from 4.7 to 8.7 (mean=6.1 and median = 6.0), and age ranged from 18 to 74 (mean = 53). Fasting interstitial glucose (FIG) was obtained via the CGM– mean FIG was 99, 112, and 139 mg/dL for the norm, PD, and T2D groups, respectively. Median FIG was 98, 111, and 137 mg/dL for the norm, PD, and T2D groups, respectively. The coefficient of determination (R^2^) between A1c and FIG was 0.375 (Figure 2A).

**Figure 2.**
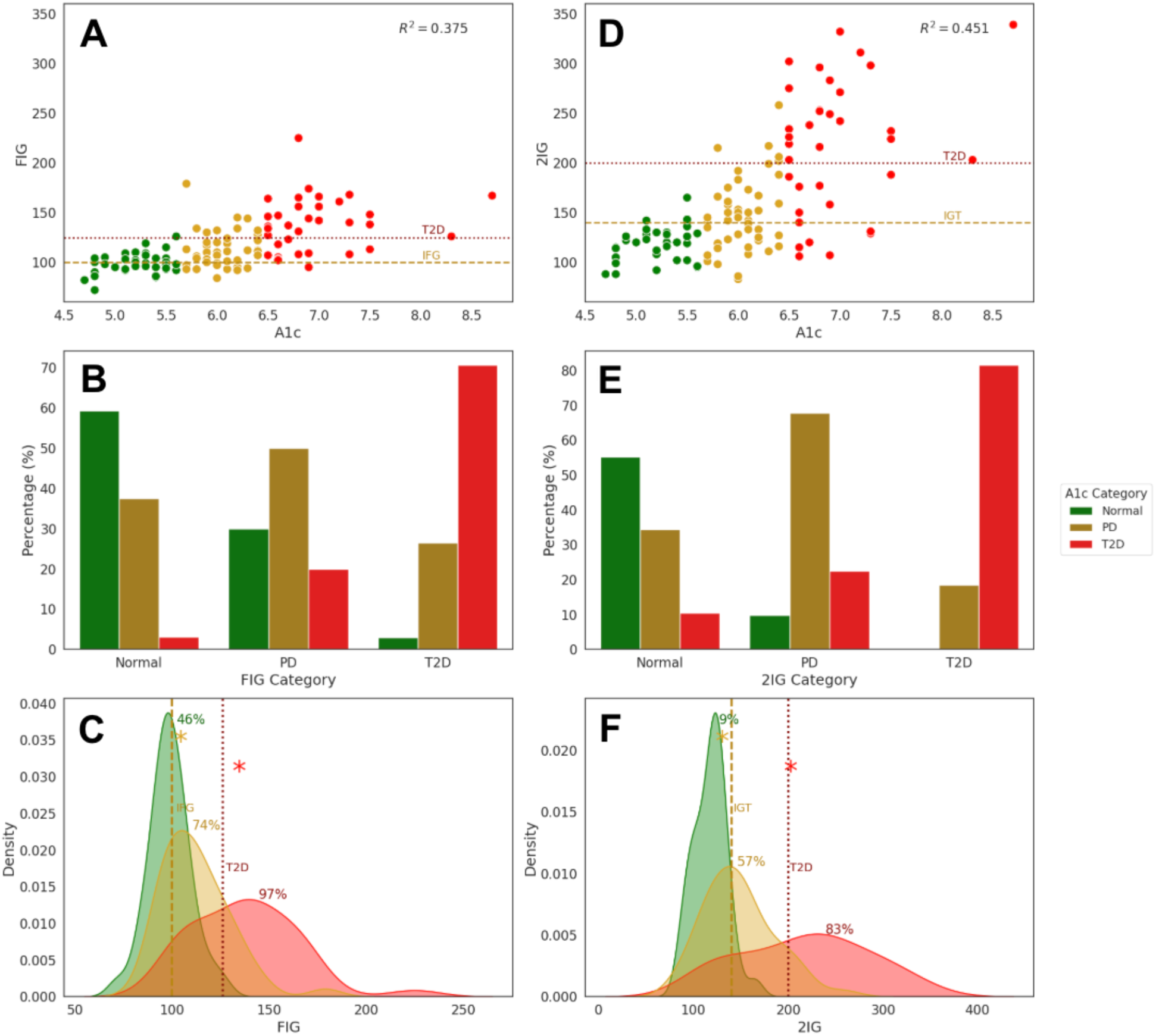
Fasting Interstitial Glucose (FIG) and 2-hour Interstitial Glucose (2IG) during the Mobile OGTT. ***(A, D)*** Scatterplots of A1c versus FIG ***(A)*** and 2IG ***(D)*** with clinical FPG-based thresholds for impaired fasting glucose (IFG; 100 mg/dL; yellow dashed line), impaired glucose tolerance (IGT; 140 mg/dL; yellow dashed line), and diabetes (125 mg/dL for FIG and 200 mg/dL for 2IG; red dotted lines). A1c-based diagnosis is denoted by the points in green, yellow, and red for the Norm, PD, and Diab groups, respectively. ***(B, E)*** Proportion of people within each A1c-based diagnostic category that are classified into the same or different categories using the clinical plasma-based diagnostic thresholds applied to CGM for either FIG- ***(B)*** or 2IG ***(E)*** on the x-axis. ***(C, F)*** Distributions of FIG (C) and 2IG (F) values displayed via kernel density estimation (KDE), divided into three curves based on A1c-based diagnosis. Vertical lines indicating the clinical plasma-based IFG and IGT (yellow dashed lines) and T2D (red dotted lines) thresholds. The yellow and red asterisks denote the optimal cutoff thresholds for PD and T2D using a grid search of values that maximally separates the A1c-defined groups, with the resulting data-driven thresholds for FIG of 104.5 mg/dL for PD (yellow) and 134.5 mg/dL for T2D (red) ***(C)***, and for 2IG: 130.5 mg/dL for PD (yellow) and 202.5 for T2D (red) ***(F)***. Annotated percentages reflect the proportion of individuals in each diagnosis group exceeding the prediabetic cutoff. 46% of A1c-defined Norm individuals are above the FIG PD cutoff, and 9% are above the 2IG PD cutoff. The yellow and red asterisk denote the optimal cutoff thresholds for PD (130.5 mg/dL) and T2D (202.5 mg/dL) respectively using a grid search of values that maximally separates the A1c-defined groups.

### Employing Fasting Plasma Glucose Diagnostic Thresholds to Aid in Clinical Interpretation of Continuous Glucose Monitor Data

Overall, most people defined as norm, PD, or T2D based on their A1c fall into the same FIG-defined class (Figure 2B). For normoglycemic and T2D, the adjacent classes were the second most populated, and the farthest classes were the least populated. Using the A1c-defined categorizations as the ground truth, FIG-based classifications have specificities of 84%, 64%, and 88% for norm, PD, or T2D, respectively, and sensitivities of 54%, 54%, and 69%.

Within the A1c-defined norm, PD, and T2D groups, 43%, 54%, and 29% of people, respectively, exhibited IFG indicative of PD based on their FIG values. IFG indicative of T2D was observed in 3%, 20%, and 69% of the A1c-defined norm, PD, and T2D groups, respectively. Nine of the people with A1c-defined PD had FIG indicative of T2D, indicating a potentially more “at-risk” phenotype for progression to T2D. Taken together, we find that IFG, defined through CGM-measured FIG, is widely prevalent in the normoglycemic group, in which 43% of A1c-based normoglycemic people met our CGM-based definition for IFG (Figure 2A, green points above lower dashed line; Figure 2C, green curve to the right of the yellow vertical dashed line) (Table 2). Notably, only one out of 35 people with A1c-based T2D had normal fasting glucose (FIG<100 mg/dl), which may be a result of changes to diet and lifestyle during the study.

### Developing Data-Driven CGM-based thresholds for Defining Impaired Fasting Glucose

Given that plasma and interstitial glucose values may diverge for a variety of reasons, it is important to assess whether data-driven thresholds for identifying IFG from CGM data are acceptable for clinical translation. Through a grid search examining a range of candidate FIG values, we determined the optimal thresholds that most accurately recapitulate A1c-based classification of glycemic status to be 104.5 mg/dL for PD and 134.5 mg/dL for T2D. These values are 4.5 mg/dL and 8.5 mg/dL higher than the corresponding FPG thresholds used clinically for classifying PD- and T2D-level IFG, indicating that CGM-based IFG classification may require higher thresholds than plasma if it is to be adopted in clinical settings for diagnosing IFG (Figure 2C). Plasma and CGM glucose measurements are known to differ (e.g., MARD>16% during OGTT in healthy persons) and CGMs tend to have higher variability and report higher glucose values than plasma, particularly at high glucose concentrations.^24–26^

Using these updated FIG thresholds yielded an overall classification accuracy of 62.93% when benchmarked against A1c-based diagnostic categories, as compared with an accuracy of 58.62% when applying traditional FPG thresholds to mOGTT data as was done previously. A modest concordance between A1c- and glucose-based classifications of glycemic status is reasonable given the known physiological and temporal differences between FIG and A1c and that discrepancies in glycemic classification based on different clinical metrics are well established ^27–29^, especially under certain conditions like dietary interventions.^30^ Improving the alignment between A1c- and CGM-based metrics, like the 4.31 percentage point increase in classification accuracy that we achieved using the data-driven FIG thresholds, could enable improved clinical decision-making by reducing diagnostic ambiguity about a patient’s glycemic state.

### Employing 2-hour Plasma Glucose Diagnostic Thresholds to Aid in Clinical Interpretation of Continuous Glucose Monitor Data

Fasting measures of glucose homeostasis provide only a partial view; the progression of dysglycemia and development of diabetes also includes a gradual rise in postprandial glucose levels.^31–34^ To explore CGM-based metrics that are corollaries to metrics used clinically to assess postprandial glucose responses, we calculated interstitial glucose values at 2 hours during a mobile OGTT (mOGTT), which we subsequently refer to as 2IG. Like the 2-hour plasma glucose measured during an OGTT, 2IG captures postprandial glucose clearance and reflects insulin-stimulated glucose uptake in peripheral tissues, particularly skeletal muscle, offering insight into β-cell function and postprandial glycemic control.^10^

Impaired Glucose Tolerance (IGT) is defined by a 2-hour serum glucose value between 140-199 mg/dl on OGTT; a 2-hour serum glucose of ≥ 200 mg/dl on OGTT is consistent with a diagnosis of T2D. Exploration of concordance between A1c-defined groupings and those based on IGT thresholds applied to 2IG on the mOGTT shows that the majority of people defined as norm, PD, or T2D based on their A1c fell into the same IGT-defined class, with the farther classes being ordinally less populated (Figure 2E. Treating A1c as a ground truth, we find that the 2IG-based classifications have specificities of 67.90%, 85.71%, and 93.83% for norm, PD, or T2D, respectively, and sensitivities of 91.43%, 45.65%, and 62.86%.

The R^2^ between A1c and 2IG in our cohort was 0.451, which is 0.076 (20.3%) higher than the R^2^ between A1c and FIG (Figure 2A, D and 4B). The mean 2IG values were 118, 148, and 217 mg/dl for the A1c-defined norm, PD, and T2D groups, respectively. The median 2IG values were 120, 144, and 224 mg/dl for the A1c-defined norm, PD, and T2D groups, respectively. The proportion of individuals falling within the IGT range on mOGTT(140 ≤ 2IG < 200) in the A1c-defined norm, PD, and T2D groups were 9%, 46%, and 20%, respectively, and those that met T2D thresholds based on 2IG levels ≥200 were 0%, 11%, and 63%, respectively (Figure 2E and F). These data suggest that, while there is a strong concordance in classification based on population means between A1c and 2IG, there is important intra-individual heterogeneity that would be missed by using only one metric.

One-fifth (7/35) of individuals with T2D based on A1c had 2IG levels that were within the range for IGT (140 ≤ 2IG < 200 mg/dL) and three-fifths (22/35) demonstrated 2IG levels consistent with diabetes (i.e.≥ 200 mg/dL). Even within CGM-based metrics, there is imperfect concordance in classifications, shown for example by the R^2^ between FIG and 2IG being 0.502 (Figure 3B), which, although higher than their individual coherence with A1c, still would result in discordance in clinical classification. Notably, 2IG showed high specificity in the normoglycemic group, with only 9% (N=3/35) of people exceeding the IGT threshold for PD and none exceeding the threshold for T2D, suggesting a largely intact postprandial insulin response despite elevated fasting interstitial glucose levels in some individuals (43% individuals showed FIG indicative of PD and 3% showed FIG indicative of T2D).

**Figure 3.**
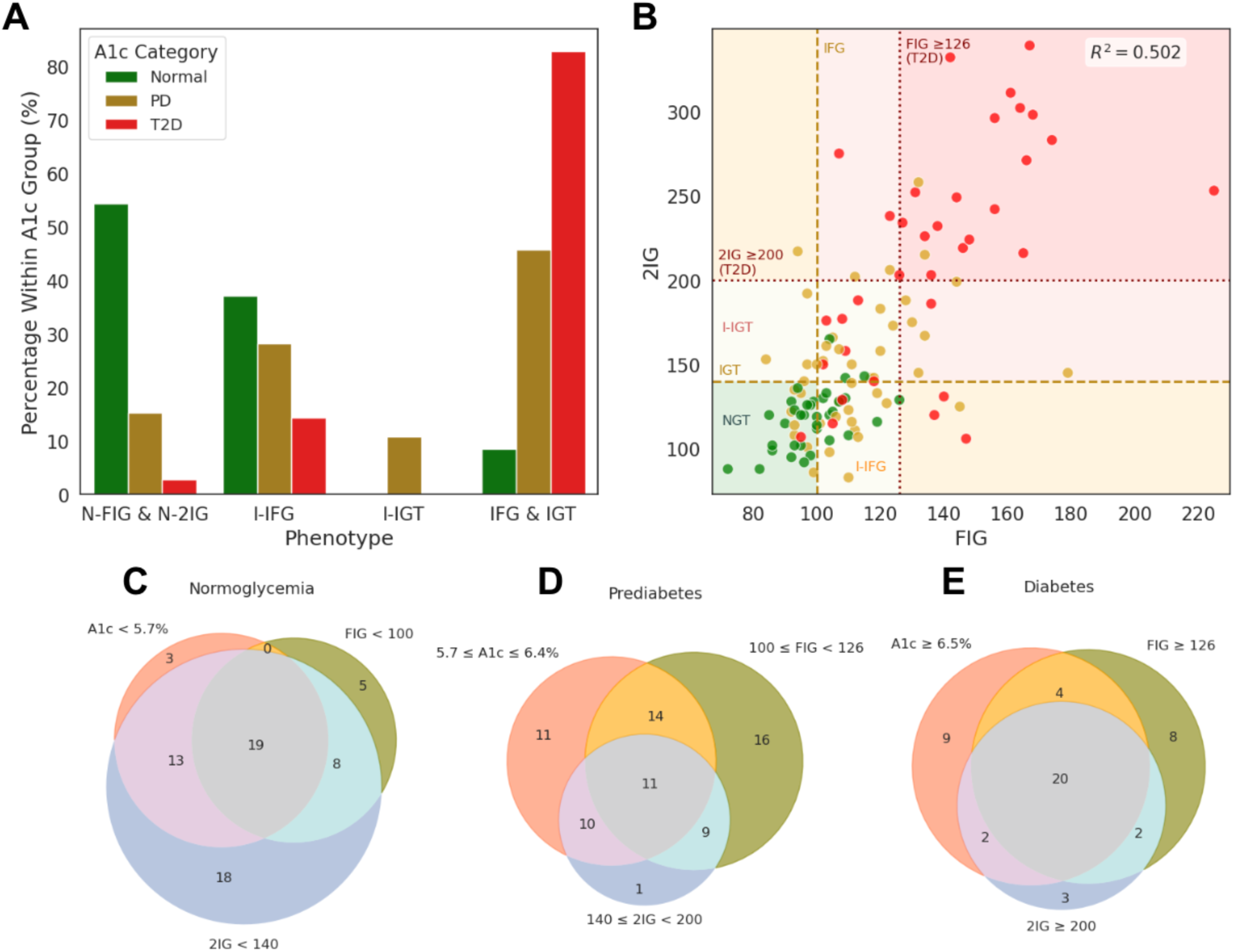
Integrating multiple metrics demonstrates concordance and discordance between mOGTT- and A1c-based classifications. ***(A)*** Distribution of OGTT-derived glucose phenotypes within A1c-defined diagnostic categories. Participants were categorized by their FIG and 2IG response into four OGTT-based groups: normal glucose tolerance (Normal FIG & 2IG), isolated impaired fasting glucose (I-IFG), isolated impaired glucose tolerance (I-IGT), and combined IFG & IGT. Bars show the percentage of individuals with each OGTT phenotype within A1c-defined diagnostic group. Notably, discordances emerge, such as individuals with normal A1c exhibiting isolated IGT or IFG, highlighting the heterogeneity of glycemic impairment. ***(B)*** Continuous relationship between FIG and 2IG with diagnostic thresholds overlaid. Each point represents a participant colored by clinical diagnosis (Norm, PD, T2D). Background shading delineates risk zones based on FIG (horizontal) and 2IG (vertical) thresholds. Color-coded risk cells illustrate diagnostic zones, with annotations for I-IFG, I-IGT, and NGT. Dashed and dotted lines mark clinical thresholds. The coefficient of determination (R² = 0.481) indicates moderate alignment between fasting and post-load glucose values, reinforcing their complementary diagnostic value. ***(C, D, E)*** Venn diagrams visualizing overlap between A1c-, FIG-, and 2IG-based classifications. Three panels separately depict the overlap of normoglycemia, PD, and Diabetes definitions based on each of the three criteria. The three Venn diagrams for T2D, PD and Norm show the number of individuals who lie within the diagnosis thresholds for that glycemic category. Each circle represents individuals meeting the corresponding threshold for that marker: A1c < 5.7% / 5.7–6.4% / ≥6.5%, FIG < 100 / 100–125 / ≥126 mg/dL, and 2IG < 140 / 140–199 / ≥200 mg/dL. While some individuals are concordantly classified across all three metrics, substantial non-overlap is observed, underscoring classification discrepancies that could impact diagnosis, treatment, and population surveillance. In the Norm group, 19 individuals meet all three normal thresholds, but substantial discordance exists: e.g., 13 participants meet only A1c and 2IG criteria but not FIG, and 8 meet A1c and FIG but not 2IG. Among PD-classified individuals, only 11 meet prediabetes thresholds across all three markers, whereas a notable number are captured by only one or two metrics (e.g., 14 by FIG and A1c alone). In the diabetes group, 18 individuals simultaneously meet the diabetic thresholds for A1c, FIG, and 2IG, but others are discordantly classified; e.g., 9 individuals meet only the A1c criterion for diabetes, while 8 meet only the FIG threshold.

### Developing Data-Driven CGM-based thresholds for Defining Impaired Glucose Tolerance

Similar to the FIG threshold analysis, we determined data-driven cut points that most accurately recapitulate A1c-based classification of glycemic status of norm, PD, and T2D. Interestingly, this analysis revealed the opposite directionality of threshold change for PD, wherein the ideal PD thresholds to be applied to mOGTT data (2IG) were actually 9.5 mg/dL *lower* than the corresponding plasma glucose thresholds used for defining IGT in clinical practice (i.e.,140-199mg/dL), and mOGTT thresholds for defining T2D were 2.5 mg/dL higher than the cut-offs used for identifying T2D by traditional OGTT (i.e., ≥ 200mg/dL). More specifically, the optimal 2IG thresholds were 130.5 mg/dL to separate normoglycemia from PD, and 202.5 mg/dL to distinguish PD from T2D (Figure 2F yellow and red asterisks, respectively). The 2IG cutoff points being lower than the typical clinical thresholds for PD is consistent with the fact that interstitial compartment glucose changes occur after blood compartment glucose changes.^35^

The data-driven 2IG thresholds yielded an overall classification accuracy of 67.24% when benchmarked against A1c-based diagnostic categories, whereas the 2-hour glucose-based thresholds applied to 2IG yielded an overall accuracy of 64.66% in classifying individuals to their A1c-based category (2.6 percentage points lower than that from our data-driven thresholds). Classification based on the 2IG threshold demonstrated modestly higher accuracy compared to FIG, which achieved an overall accuracy of 58.62%. This aligns with prior findings suggesting that 2-hour OGTT measurements offer greater diagnostic value for identifying both T2D and PD than FPG.

### Diabetes Risk Stratification using CGM-based Measures of IGT and IFG

Given that FIG and 2IG each have three clinical categorizations (norm, PD, T2D), there are nine possible states, or profiles, that an individual may have, and each confers a different level of risk of glycemic deterioration. IFG and IGT reflect intermediate disruptions in glucose metabolism resulting from distinct pathophysiological mechanisms that each precede the onset of T2D. Isolated IFG and isolated IGT are defined as impaired fasting glucose levels in the absence of IGT (I-IFG: 100 ≤ FIG < 126 mg/dL) (Figure 3B, middle vertical shaded area), and impaired glucose tolerance (I-IGT: 140 ≤ 2IG < 200 mg/dL) with normal fasting glucose levels) (Figure 3B, middle horizontal shaded area), respectively.^36–39^ The presence or absence of IFG and/or IGT has important clinical implications, as they confer different risks related to progression to diabetes. While IFG may represent a more prevalent phenotype of PD ^40^, individuals with isolated IFG tend to progress to T2D more slowly than those with isolated IGT. The joint presence of IFG and IGT represents a higher risk than the presence of either state in isolation. ^31,41–43^

Using these definitions, 12 (34.3%), 12 (26.1%), and 2 (5.7%) individuals from the norm, PD, and T2D categories, respectively, displayed I-IFG (Figure 3A, and the bottom trisection of the middle vertical shaded region of Figure 3B, marked I-IFG). Surprisingly, no one in the normoglycemic or T2D groups showed PD-level I-IGT, while 4 people in PD showed PD-level I-IGT. The presence of both IGT and IFG is the highest risk state, and of the norm, PD, and T2D groups, 3/35 (8.6%), 11/46 (23.9%), and 6/35 (17.1%) of people showed combined IGT and IFG, respectively (i.e., 100 ≤ FIG < 126 and 140 ≤ 2IG < 200). Overall, 27 individuals showed NGT (FIG<100 mg/dL and 2IG <140 mg/dL), represented in the green shaded region on the bottom left of Figure 3B (marked NGT).

Similarly, 13 (37%), 13 (28%), and 5 (14%) individuals from the norm, PD, and T2D categories respectively displayed I-IFG, including IFG indicative of T2D (I-IFG+T2D) (T2D+IFG: 100 ≤ FPG mg/dL). In the PD group, 5 (11%) people showed isolated IGT, including IGT indicative of T2D (I-IGT+T2D) (IGT: 140 ≤ 2IG mg/dL) with no one in normoglycemic or T2D groups showing I-IGT+T2D. From the norm, PD, and T2D groups, 3(9%), 21 (46%), and 29 (83%) of people showed combined IGT and IFG (i.e., FPG: ≥ 100 mg/dL and 2IG ≥ 140 mg/dL).

### Discrepancies in Glycemic Classification by OGTT and A1c Reveal Subclinical Heterogeneity

Building on these findings, we next examined how standard diagnostic markers (A1c, FPG, 2-hour OGTT glucose) and CGM-based proxies– FIG and 2IG– align or diverge in classifying individuals by their glycemic status. Across A1c, FIG, and 2IG, 50/116 total people (43%) were consistently classified by all three metrics into either the norm, PD, or T2D categories (19/35 (54%) normoglycemic (Figure 3C), 11/46 (24%) PD (Figure 3D), and 20/35 (57%) T2D (Figure 3E)). In other words, 66/116 total people had discrepancies in classification depending on the metric used, which is consistent with literature exploring concordance between A1c, FPG, and 2-hour OGTT-based diagnoses.^5,28,44^

More specifically, looking at individuals who had A1c values below the diagnostic threshold for T2D, 5/81 and 10/81 exhibited 2IG and FIG levels consistent with T2D, respectively. Of those with A1c values below the diagnostic threshold for PD, 3 and 15 had 2IG and FIG levels consistent with PD, respectively. These findings are consistent with existing clinical dialogue that CGM provides more sensitive biomarkers for early dysglycemia than A1c, especially in high-risk groups, such as those with impaired glucose tolerance and impaired fasting glucose that has not yet translated to an A1c in the PD/T2D range.

Conversely, of the 35 people classified as T2D by their A1c, 13 had 2IG values below the T2D cutoff. Of the 46 classified as PD, 20 had 2IG values below the PD cutoff. This could imply a more reversible or ‘recently transitioned’ phenotype. Because our study was conducted remotely with limited supervision, we also cannot exclude the possibility of suboptimal adherence to the mOGTT protocol that may have blunted the glycemic response (e.g. participant may not have finished the beverages within the specified time period).

For those 46 people classified as PD, 25 individuals had 2IG outside the PD range, (with 5/46 showing 2IG higher than the PD threshold), while ten individuals had elevated 2IG suggestive of PD, but A1c levels outside the PD criteria (7 showing A1c indicative of T2D and 3 with A1c indicative of normoglycemia). Overall, 25 had FIG values within the range for PD diagnosis and did not have a diagnosis of PD from A1c. This suggests the potential presence of early impaired insulin sensitivity (especially hepatic), with or without IGT; Identification of this condition would enable early monitoring to prevent progression.^45,46^ Since OGTT is rarely performed in individuals with normal A1c (outside of pregnancy), at-home mOGTT has the potential to serve as a personalized and convenient risk tracking tool that allows users to identify early patterns of dysglycemia, empowering them to enact lifestyle changes before they progress to diabetes. mOGTT provides complete interstitial glucose curves, facilitating data-driven mapping between curve shape features and underlying metabolic subphenotypes.

### Characterization of mOGTT Curves Across the Glycemic Spectrum

OGTT responses increased in intensity of glucose response amplitude from normoglycemic to PD to T2D (Figure 4A).^1^ While plasma glucose thresholds for OGTT-based diagnosis are well-established, no equivalent CGM-based thresholds exist to date. The traditional plasma-based thresholds for the OGTT curves (beyond FPG and 2-hour glucose) include peak glucose concentration of 140 mg/dL and peak onset timing of 60 minutes – values below these thresholds are considered normal.^10^ Here, the equivalent data-driven thresholds for interstitial glucose, as measured by CGM, were found to be as follows: peak glucose concentration of 152.5 mg/dL and peak onset timing of 38 minutes.

**Figure 4:**
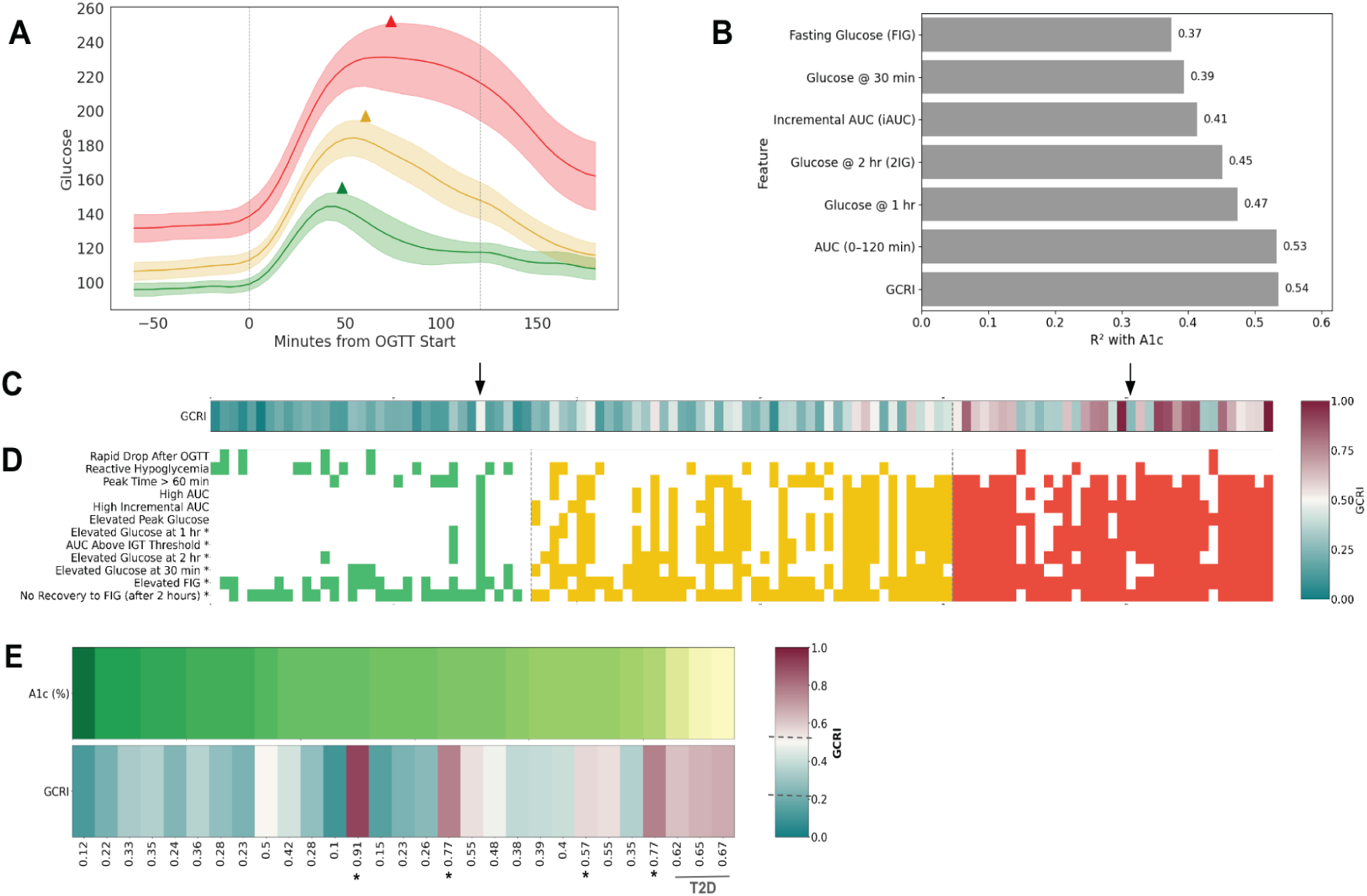
Temporal, correlational, and phenotypic characterization of glycemic responses. ***(A)*** Mean glucose response curves over OGTT time with 95% confidence intervals stratified by diagnostic group. Glucose values were aligned to a 5-minute grid for each participant and averaged across individuals within each group (Norm, PD, T2D). Diagnostic groups show distinct glucose trajectories. Triangular markers denote each group’s average time-to-peak and corresponding peak glucose value, revealing delayed and elevated peaks in PD and T2D. ***(B)*** Explained variance (R²) between OGTT-derived glucose metrics and A1c. Linear regression was used to quantify the strength of association between individual OGTT time-point or composite metrics (e.g., iAUC, 2IG) and A1c. GCRI exhibited the strongest explanatory power among all mOGTT metrics. ***(C)*** Participant-level dysglycemia subphenotypes and GCRI scores ordered by A1c. Each column represents one participant, sorted by A1c in ascending order. Rows show presence/absence of phenotypic dysglycemia features (e.g., delayed peak, reactive hypoglycemia, no recovery) and are color-coded by diagnostic group and the features used to compute GCRI are denoted by ‘*’. The top heatmap encodes the GCRI, normalized across participants. GCRI is calculated for each participant based on mOGTT metric deviations (ranging from 0 to 1, where 0 represents the healthiest mOGTT profile and 1 represents the presence of the most severe dysglycemia based on mOGTT). Vertical dashed lines indicate A1c thresholds for PD and T2D. **(D)** External validation cohort: A1c vs. GCRI spectra (sorted by A1c). Each column is one participant (x-axis labels show GCRI). The top row encodes A1c values using the green (Norm), yellow (PD), red (T2D) colormap, normalized to a fixed A1c range of 4.5–9.5%. The bottom row encodes the GCRI, displayed on a fixed [0, 1] scale (higher risk = red), using the muted green (low GCRI), yellow (medium GCRI), and red (high GCRI) color scheme to allow visual comparison across panels. Participants are ordered by A1c (ascending) so correspondence between A1c and composite risk can be visually assessed column-by-column. The color bar legend indicates the mapping for the GCRI scale.

In T2D, the mean peak interstitial glucose concentration was 98 and 56 mg/dL higher than in normoglycemia and PD, respectively. The absolute peak values were 253 mg/dL for people with T2D, as compared with 197 mg/dL and 155 mg/dL for people with PD and normoglycemia. The T2D and PD groups also exhibited delayed glucose peaks at 74 and 60 minutes, respectively, which is indicative of postprandial glucose response dysregulation. For comparison, the normoglycemic group peaked at 48 minutes. The highest resolution OGTT sampling to date comparing norm, PD, and T2D characteristics has been done using blood glucose– this study showed that in normoglycemic, insulin-sensitive individuals, the time to glucose peak typically occurs at or before 30 minutes, and a later time to peak (≥ 60 minutes) occurs frequently in adults with type 2 diabetes, but the sampling resolution was still relatively coarse.^18^ The later time observed in our study is also likely due to differences between blood and interstitial glucose dynamics. More people in the PD and T2D categories did not return to baseline within 120 minutes (38 and 32 individuals, respectively) than in the normoglycemic group (23 individuals). The mean time to baseline was 106, 115, and 116 minutes for norm, PD, and T2D respectively. This is consistent with existing literature that describes expected return to baseline for individuals with normoglycemia within 2-3 hours of glucose ingestion.^10^

Together, these findings suggest delayed glucose recovery in PD and T2D relative to normoglycemia. Interpretation of the mOGTT responses using CGM, however, must account for the documented 5–25-minute lag between interstitial and blood glucose postprandially due to both blood-to-interstitial glucose transport and instrument-related delay (this lag is only 5-6 minutes in an overnight fasted state in healthy adults).^35,47^ Such differences in technology underscore the need to establish normative mOGTT values to enable CGM-based diagnostics and monitoring in ambulatory settings. The lag also suggests that extending mOGTT duration to 3-4 hours, particularly in PD and T2D, may be important to capture the full glucose uptake and recovery dynamics.

### CGM-based Metrics of mOGTT Responses

In the clinic, OGTT usually involves point measurements, often only at baseline and either 1-or 2-hours post-ingestion, but sometimes 30- minutes or 3 hours post-ingestion, and less frequently at multiple post-ingestion time points. These point measurements are then used to calculate additional informative metrics: the area under the curve (AUC), and the incremental AUC (iAUC).^10,17,48–50^ We evaluated the six commonly used OGTT metrics from the mOGTT data in the DiabetesWatch cohort: fasting plasma glucose (FPG), 30-minute, 1-hour, and 2-hour glucose, along with AUC and iAUC from the 2-hour mOGTT window.

The aggregate measures of all six metrics followed expected clinical gradients– highest in the T2D group (Figure 4A, red), intermediate in PD (Figure 4A, yellow) and lowest in normoglycemic (Figure 4A, green) (Table 2), demonstrating that mOGTT replicates clinically meaningful patterns.^10^

Among all metrics, FIG exhibited the greatest proportion of individuals exceeding the impaired fasting glucose (IFG) threshold across all groups in the DiabetesWatch population, out of all 6 metrics. It showed the highest sensitivity in the T2D group and the lowest specificity in the normoglycemic group, when A1c was used as the reference diagnostic. (Table 2 and Figure 2B).

None of the six metrics could distinguish between diagnostic groups with 100% accuracy (Table 1). FIG, 2-hour glucose, and A1c are the most common metrics used clinically for assessing T2D and PD, and there is known diagnostic discordance between them ^28,29^. We explored whether there might be comparatively less discordance between the mOGTT metrics in free-living settings. Tucker et al. previously reported, for T2D, a concordance of only 34% between OGTT 2-hour glucose and A1c, and 44% between 2-hour glucose and FPG. For PD, the concordances were and 46% and 63%, respectively, and for norm, they were 81% and 65%, respectively.^28^

**Table 1.** Demographics of DiabetesWatch Study Population.

|  | Euglycemia | Prediabetes | Type 2 Diabetes | Total |
| --- | --- | --- | --- | --- |
| Female | 18 | 25 | 20 | 62 |
| Male | 17 | 21 | 15 | 54 |
| White | 23 | 24 | 19 | 66 |
| African American | 4 | 14 | 12 | 30 |
| More than one race | 5 | 4 | 2 | 11 |
| Asian | 2 | 3 | 2 | 7 |
| Amer Indian/Alaska Nat | 0 | 1 | 0 | 1 |
| Unknown Race | 1 | 0 | 0 | 1 |
| Hispanic | 12 | 9 | 2 | 23 |
| **Total** | **35** | **46** | **35** | **116** |

**Table 2.** OGTT Metrics Overall and by Diagnosis Category.

| **Metric** | **Statistic** | **Total** | **Normoglycemic** | **Prediabetes** | **Type 2 Diabetes** |
| --- | --- | --- | --- | --- | --- |
| FIG | Elevated FIG ($\geq 100$) N (%) | 84 (72.4%) | 16 (45.7%) | 34 (73.9%) | 34 (97.1%) |
| FIG | Mean ± SD | 116.0 ± 24.8 | 98.7 ± 10.6 | 112.5 ± 17.7 | 137.9 ± 27.1 |
| FIG | Median (range) | 109.0 (72.0–225.0) | 98.0 (72.0–126.0) | 110.5 (84.0–179.0) | 137.0 (95.0–225.0) |
| FIG | Correlation with GMI | 0.83 | 0.88 | 0.62 | 0.74 |
| Thirty-Min Glu | Elevated 30 min glu ($\geq 165.6$) N (%) | 49 (42.2%) | 5 (14.3%) | 17 (37.0%) | 27 (77.1%) |
| Thirty-Min Glu | Mean ± SD | 164.1 ± 35.5 | 136.7 ± 22.3 | 162.4 ± 26.5 | 193.9 ± 33.8 |
| Thirty-Min Glu | Median (range) | 158.5 (101.0–284.0) | 136.0 (101.0–187.0) | 158.5 (107.0–232.0) | 196.0 (133.0–284.0) |
| Thirty-Min Glu | Correlation with GMI | 0.72 | 0.57 | 0.43 | 0.60 |
| One-Hr Glu | Elevated 1 hour ($\geq 155$) glu N (%) | 77 (66.4%) | 8 (22.9%) | 36 (78.3%) | 33 (94.3%) |
| One-Hr Glu | Mean ± SD | 183.5 ± 54.6 | 135.9 ± 34.1 | 183.7 ± 37.4 | 230.8 ± 49.4 |
| One-Hr Glu | Median (range) | 176.5 (83.0–328.0) | 135.0 (83.0–250.0) | 184.0 (85.0–250.0) | 236.0 (112.0–328.0) |
| One-Hr<br>Glu | Correlation with<br>GMI | 0.78 | 0.49 | 0.58 | 0.70 |
| Two-Hr<br>Glu | Elevated 2 hour<br>($\geq 140$) glu N<br>(%) | 58 (50.0%) | 3 (8.6%) | 26 (56.5%) | 29 (82.9%) |
| Two-Hr<br>Glu | Mean ± SD | 159.6 ± 59.3 | 117.8 ± 17.0 | 148.1 ± 37.3 | 216.6 ± 66.8 |
| Two-Hr<br>Glu | Median (range) | 139.5 (83.0–<br>339.0) | 120.0 (88.0–165.0) | 143.5 (83.0–<br>258.0) | 224.0<br>(106.0–<br>339.0) |
| Two-Hr<br>Glu | Correlation with<br>GMI | 0.82 | 0.56 | 0.55 | 0.74 |
| AUC | Elevated AUC<br>($\geq 305 \times 60$) N<br>(%) | 58 (50.0%) | 2 (5.7%) | 25 (54.3%) | 31 (88.6%) |
| AUC | Mean ± SD | 19167.0 ±<br>5332.9 | 14549.6 ± 2389.4 | 18694.7 ±<br>3240.2 | 24405.3 ±<br>5108.1 |
| AUC | Median (range) | 18318.8<br>(10547.5–<br>34292.5) | 14285.0 (10547.5–<br>23425.0) | 18711.2<br>(12115.0–<br>25580.0) | 24280.0<br>(16275.5–<br>34292.5) |
| AUC | Correlation with<br>GMI | 0.85 | 0.67 | 0.64 | 0.77 |
| iAUC | Mean ± SD | 5811.7 ±<br>3279.3 | 3216.8 ± 1764.1 | 5737.3 ±<br>2582.1 | 8504.4 ±<br>3167.6 |
| iAUC | Median (range) | 5290.0<br>(595.0–<br>14517.5) | 3038.0 (1350.0–<br>11465.0) | 5712.5 (595.0–<br>12007.5) | 8787.5<br>(1754.0–<br>14517.5) |
| iAUC | Correlation with<br>GMI | 0.65 | 0.27 | 0.31 | 0.52 |

**Table 3.**
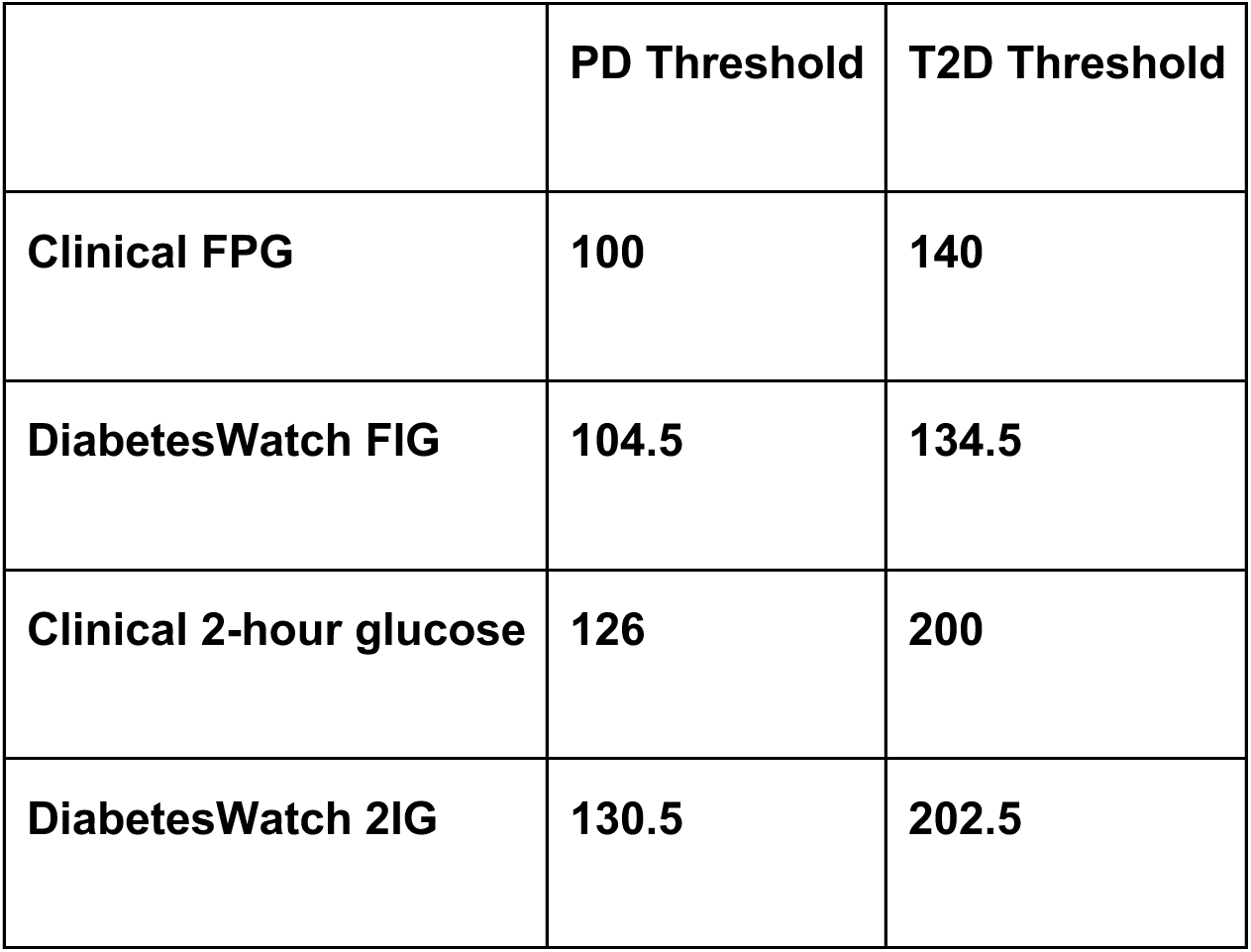
Clinical vs. data-driven dysglycemia thresholds for FPG and 2-hour OGTT glucose.

In contrast, in the DiabetesWatch cohort, 2IG-based classification matched A1c-based classification in 82% of T2D participants and FIG–based classification in 82%; the corresponding proportions were 68% (A1c) and 65% (FIG) in PD, and 55% (A1c) and 47% (FIG) in normoglycemia, all based on the clinical OGTT and A1c diagnostic threshold. While these findings suggest improved alignment in free-living settings for PD and T2D, partial overlap between diagnostic criteria persists, consistent with existing literature. ^51,52^

Of post-glucose-load mOGTT metrics, 1-hour glucose emerged as the most sensitive, with 8/35 (22.9%), 35/46 (76.1%), and 33/35 (94.3%) of individuals from the normoglycemic, PD, and T2D categories, respectively, demonstrating 1-hour glucose elevations above the IGT threshold. This is consistent with recent literature that suggests that 1-hour post-load plasma glucose is a more sensitive and practical method to screen for early dysglycemia and T2D in people at risk. ^7^

To further contextualize the six OGTT-derived metrics within the broader landscape of glycemic health, we examined their relationship with the glucose management indicator (GMI), calculated from 13 days of CGM data (excluding first 24 hours of CGM data to avoid any calibration-related inaccuracies) using the iglu_py package in Python.^53,54^ Across all groups, AUC showed the strongest correlation with GMI among all six OGTT metrics as measured by Pearson correlation. Overall, GMI correlations were highest in the T2D cohort for all metrics, except FIG, where the correlation was strongest in the normoglycemic group at 0.89. These results suggest that CGM-derived OGTTs not only reflect short-term glycemic responses but are also predictive of longer-term glycemic health states.

In examining the relationship between each of the six OGTT metrics and A1c (Figure 4B), all showed different levels of shared information content, with AUC having the highest correlation with A1c at R^2^ = 0.53. This suggests the existence of non-redundant information contained within the individual OGTT metrics, indicating that together they may provide a more comprehensive view of glycemic health. Given this and the prior results showing an inability of existing metrics to clearly discern subphenotypes, we next introduce a complementary evaluation tool, the “Glucose Challenge Response Index (GCRI)” to improve discernment in glycemic subphenotypes as well as enable insights on shorter time scales than A1c allows.

### Proposing a Composite mOGTT Score for Subphenotyping

To determine whether the six mOGTT metrics could be combined to generate a single informative and quantifiable phenotype to capture and summarize multiple dimensions (e.g., magnitude, pattern) of glycemic dysfunction, we created a composite mOGTT score called GCRI ranging in value from 0 (“healthiest”) to 1 (“most dysfunction”) as the normalized sum of the distances between the mOGTT metrics and their corresponding diagnostic thresholds. The GCRI is calculated by centering each mOGTT metric on its clinical threshold (e.g., for FIG the PD threshold is 100 mg/dL), normalizing by the standard deviation, averaging across FIG, 30IG, 1IG, 2IG, AUC, and end-start difference for each person, and then rescaling. With R^2^=0.54, the GCRI had the highest correlation with A1c as compared with existing mOGTT metrics (Figure 4B), demonstrating its relatively higher information content.

On average, individuals with A1c-defined T2D had the highest GCRI scores (median = 0.62), the widest range [0.26, 1.00], and the highest variance (SD = 0.23). PD showed intermediate GCRI scores (median = 0.33; range [0.08, 0.61]; SD = 0.13). Normoglycemic individuals had the lowest median (0.17), tightest range [0, 0.48], and lowest variance (SD = 0.09). These results demonstrate the increasing heterogeneity in glycemic regulation with worsening metabolic status and generate an intuitive stratification by relative level of dysglycemia (Figure 4C). The optimized thresholds for the GCRI were 0.22 for PD and 0.56 for T2D.

While the GCRI summarizes overall risk, visualization of the mOGTT metrics that serve as underlying components of the index, and comparison with other CGM-based indicators (e.g., whether or not a person returns to baseline), reveal which domains (e.g., fasting, early peak, late recovery, total exposure) drive risk and where an individual lies along the dysglycemia spectrum (Figure 4D). This enables a more nuanced understanding of why a given individual’s GCRI is high or low. For example, the participant with the lowest GCRI (0) exhibits the fewest dysglycemia flags– only experiencing “no recovery to FIG”– illustrating how a low global risk can still harbor a specific and subclinical metabolic abnormality.

#### Case Studies

An illustrative case highlighted in Figure 4C (left black arrow) shows an individual classified as normoglycemic based on their A1c who exhibited a high GCRI (0.48) which would classify them as PD by the mOGTT thresholds, aligning them more closely with PD and borderline T2D mOGTT profiles. Despite their normal A1c value, this participant displays abnormalities like elevated 2IG, elevated FIG, and delayed time-to-peak glucose– substantial evidence of dysglycemia that would be missed through current clinical methods relying on A1c. This A1c-mOGTT discordance underscores a well-recognized limitation of A1c: as a three-month average, it can miss early or intermittent dysglycemia. In contrast, the GCRI integrates contemporaneous glucose dynamics, offering insight into the pathophysiology of progressive dysglycemia. Taken together, these findings suggest that the composite score can flag early metabolic risk and motivate closer clinical follow-up or earlier intervention even when A1c remains within the normative range.

In contrast to the participant who had hidden dysglycemia, we also observed one A1c-defined T2D participant with a low GCRI (0.26), near the norm/PD GCRI threshold (Figure 4C, right black arrow). The accompanying heatmap showed few dysglycemic features overall. A plausible interpretation is recent improvement in glycemic physiology (e.g., lifestyle changes) that is captured promptly by the GCRI but which would take longer to register on A1c, given its multi-month averaging window. For individuals actively working to reverse T2D, the composite GCRI thus provides a more responsive indicator for tracking progress. Together with the prior discordant case on the other end of the spectrum, this example highlights the complementary value of the GCRI for risk stratification and ongoing clinical monitoring beyond A1c.

Across the entire cohort, “no recovery to baseline FIG” emerges as the most prevalent dysfunction (93/116, 80% of the cohort), followed by elevated fasting glucose (84/116, 72%). Notably, elevated fasting glucose in isolation (i.e. elevated FIG without other dysglycemia indicators shown in the heatmap in fig 4d) occurs in the norm (2/35, 5.7%) and PD (3/46, 65%) groups but never in T2D (0/35, 0%) where abnormalities typically co-occurred.

#### Dysglycemia classification based on GCRI

We next evaluated whether the GCRI could better distinguish between A1c-defined diagnostic groups as compared with the mOGTT metrics (FIG, 2IG) explored earlier. The GCRI achieved an overall classification accuracy of 72%, which was higher than FIG (59%) and 2IG (65%). Per-class accuracy was highest for norm (77%), followed by PD (76%) and T2D (63%). Most misclassifications involved T2D assigned to the PD category, and importantly, there were no misclassifications between the normoglycemic and T2D categories, while such “extreme” misclassifications did occur using single mOGTT metrics. Specifically, 27/35 norm (77%), 35/46 PD (76%), and 22/35 T2D (63%) were correctly classified. Of those with T2D, 13 were misclassified as PD. The GCRI demonstrated a sensitivity of 77%, 76% and 63% in the norm, PD, and T2D groups respectively and specificities of 89%, 70% and 98% in the norm, PD and T2D groups respectively. Overall, the GCRI provided a more even accuracy profile across all three categories, while FIG favored detection of T2D at the expense of norm, and 2IG favored separation of normoglycemic from T2D but showed weaker performance for PD (Supplemental Figure 1). Ultimately, the GCRI demonstrated superior performance to any single mOGTT metric alone.

#### External Validation of GCRI

To evaluate its generalizability and utility for glycemic subphenotyping, we implemented the GCRI in an external dataset comprising primarily individuals without T2D, but who had metabolic dysregulation that could be characterized by between one and four metabolic dysfunction hallmarks (Muscle insulin resistance, Hepatic insulin resistance, β cell dysfunction, and Incretin effect) (N=29; norm=16, PD=10, T2D=3; A1c 5.5-7); Figure 4E). ^19^ This cohort exhibited more pronounced dysglycemia within the normoglycemic and prediabetic ranges as compared with the DiabetesWatch cohort and had an intuitively weaker concordance between mOGTT metrics and A1c (R²=0.35), FIG (R²=0.36), 1-hour glucose (R²=0.26), and 2IG (R²=0.32). Leveraging available disposition index (DI) measurements, which is a measure of β-cell function independent of insulin resistance, we evaluated the performance of the mOGTT composite score for detecting metabolic dysfunction. ^19,55^ DI in this cohort ranged from 0.54 to 6.58, with a median DI of 1.64. Complete β-cell compensatory dysfunction was considered to be DI < 1.2, intermediate dysfunction was 1.2 ≤ DI ≤ 2.2, and normal β-cell function was DI > 2.2.

The GCRI range [0.1, 0.91] lay fully within the DiabetesWatch range [0, 1], indicating that the DiabetesWatch dataset effectively captures OGTT dynamics across a wide range of severity. The GCRI range and median value in the external dataset’s normoglycemic and PD groups were wider and higher, respectively, than in the counterpart measures from the DiabetesWatch dataset. Specifically, in the normoglycemic group, the external dataset’s range was [0.10, 0.91] and the median was 0.27, while in DiabetesWatch, the range was [0, 0.48] and the median was 0.17. Similarly, the external dataset’s PD group range was [0.35, 0.77] with median 0.51, while DiabetesWatch was [0.08, 0.61] with a median score of 0.33. These distributional differences are likely due to the external dataset’s study design enriching for metabolic dysfunction.

Of the three individuals in the external dataset who had T2D-level A1c, all had GCRI above the T2D threshold derived from the DiabetesWatch dataset, validating the ability of the GCRI to discern more extreme diabetes phenotypes (Figure 4E, right side). Overall, four individuals had GCRIs above the T2D threshold derived from the DiabetesWatch dataset but A1c below the T2D threshold (Figure 4E, asterisks), all of whom had DI-related dysfunction. The GCRI detected masked dysglycemia in three participants; For example, participant S103 had an A1c of 5.5, well within the normoglycemic range, but GCRI of 0.91, and subphenotyping revealed this participant to have clinically-validated beta cell dysfunction (DI of 0.97) and insulin resistance (SSPG=267 mg/dL). Similarly, two individuals with A1c in the PD range had mOGTT values higher than the GCRI for all three T2D individuals at a GCRI of 0.77 each, and both demonstrated insulin resistance with SSPG of 225 mg/dL and 207 mg/dL, and β-cell dysfunction with DI of 1.1 and 0.6. Further, both participants showed FIG and 2IG in the T2D range and 1-hour glucose, which were well above the T2D threshold. More detailed exploration of the relationship between the GCRI and DI showed a higher correlation (R^2^ = 0.29) as compared to FIG vs DI (R^2^ = 0.14) and 2IG vs DI (R^2^ = 0.24), highlighting the value of the mOGTT for detecting early signs of dysglycemia.

### Data-driven groupings that enable intermediate risk assessment

While the GCRI acts as a convenient and useful metric that takes on a single continuous value along a spectrum, classifications tend to be more clinically useful. Further, the GCRI does not give underlying information about *which* features are abnormal and *by how much*. Thus, we pursued an alternative method of assessing relative dysglycemia within the DiabetesWatch population by creating groupings in the original six-dimensional space made up of the constituent mOGTT features and compared these groupings to the A1c-based groupings. This analysis revealed three groups which could be approximately identified as 1) Glucose Tolerant (N=68), 2) Glucose Intolerant (N = 36), and 3) Severely Glucose Intolerant (N=12). These groups demonstrated higher between-group separation (average between-cluster variance = 234.8), and higher within-group similarity (average within-cluster variance = 2.0; Silhouette score = 0.41; Calinski–Harabasz index = 117; weighted purity = 0.662) as compared with the A1c-defined groups (average between-cluster variance = 145.31; average within-cluster variance = 3.59; Silhouette score = 0.096; Calinski-Harabasz index = 40.51; weighted purity = 0.461) (Figure 5A; Suppl. Figure 2). In all, the mOGTT feature-based groupings were substantially tighter (more homogeneous) than the A1c-based groupings.

**Figure 5.**
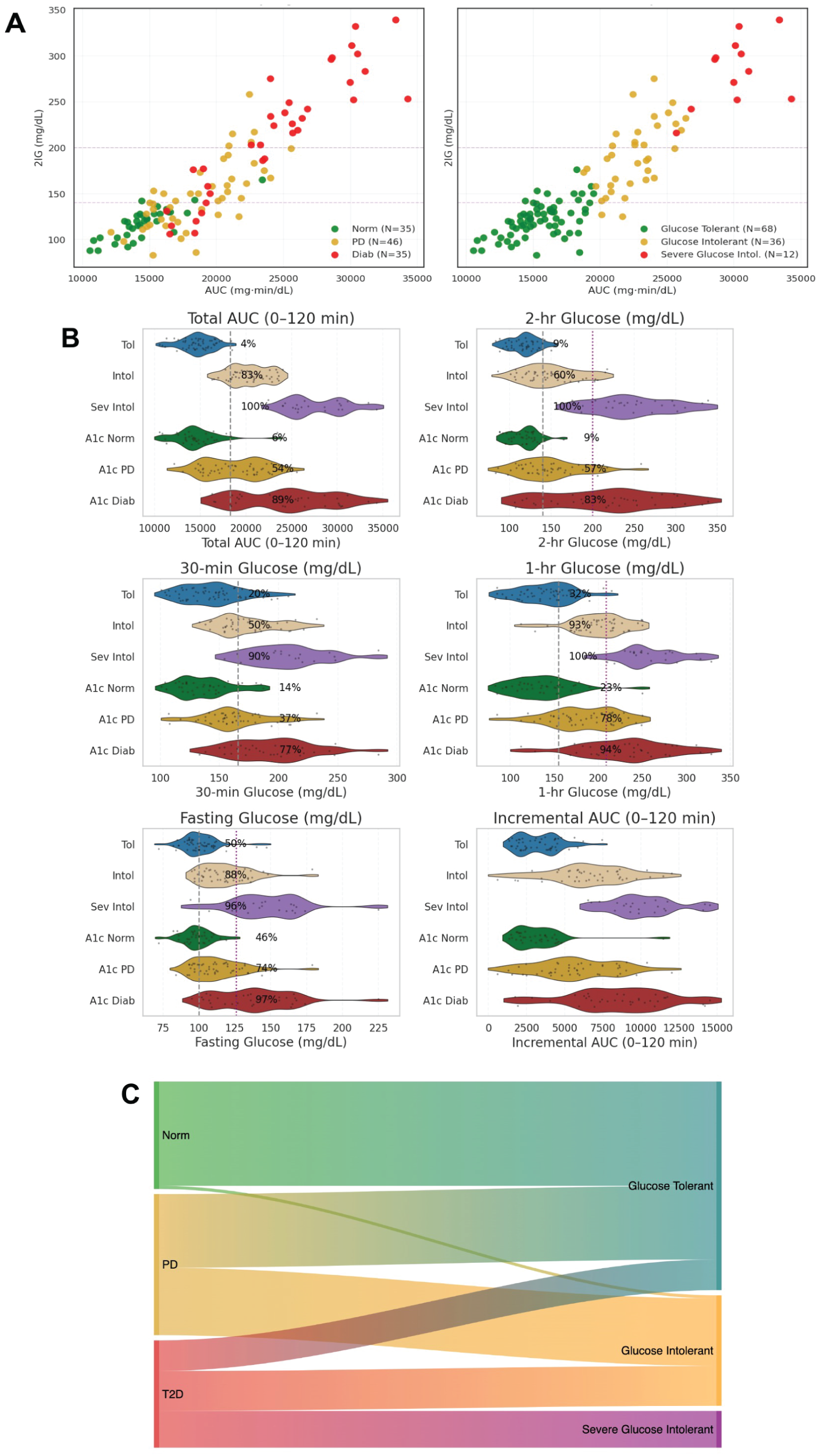
Data-driven clustering of mOGTT responses. ***(A)*** AUC and 2IG distribution by A1c and data-driven cluster ***(B)*** Distribution of 6 OGTT metrics by mOGTT LOMO model cluster-based categories in the top half and A1c-diagnosis categories in the bottom half of each metric’s panel. The vertical dashed grey and purple lines denote the clinical PD and T2D threshold values respectively for each metric (where applicable). The cluster-based categories are shown in blue (corresponding to glucose tolerant or tol cluster), orange (corresponding to intermediate dysglycemia or intol cluster) and purple (corresponding to severe dysglycemia or sev intol cluster), while the A1c-diagnosis categories are shown in green (normoglycemia), yellow (PD) and red (T2D). The annotations show the % of individuals within that group or cluster that showed a value outside the diagnostic threshold for PD ***(C)*** Proportion of participants in each A1c-based diagnostic cluster on the left (colors green, yellow, and red corresponding to normoglycemia, PD, and T2D groups respectively) that were classified into the respective mOGTT-based cluster on the right (with colors blue, orange, and purple corresponding to glucose tolerant, intermediate dysglycemia, and severe dysglycemia clusters respectively). Stream width is proportional to the number of participants in each transition. Most normoglycemic individuals were classified into the glucose tolerant cluster while one normoglycemic individual was classified into the intermediate dysglycemia cluster. From the PD group, 24 and 22 individuals were classified in the glucose tolerant and intermediate dysglycemia clusters respectively. In the T2D group, 10, 13, and 12 participants were mapped to the glucose-tolerant, intermediate-dysglycemia, and severe dysglycemia clusters, respectively.

This improved separability between groups is well visualized through the group-wise distributional differences for each of the individual mOGTT features (Figure 5B). For example, AUC has 83.6% overlap in the A1c-based groupings but only 8.6% overlap in the data-driven groupings. Such separations persist even when that metric is held out of the clustering algorithm (Leave-One-Metric-Out, or LOMO), demonstrating the robustness of the groupings and their clinical meaningfulness. Across all LOMO models, purity remained consistently high, peaking when 30-min glucose was omitted (overall purity = 0.713), followed by FIG (0.700). AUC and 2IG were the strongest contributors to internally tight, physiologically coherent clusters; removing AUC produces the largest decline in overall purity, while removing 30-min glucose improves homogeneity relative to the full model, suggesting that the early timepoint may add noise or create overlaps in phenotypes already captured more cleanly by later OGTT measures. LOMO supports the added value of integrating the multiple metrics from the full mOGTT window and justifies using the GCRI, which has the same underlying features, for risk stratification in real-world settings.

Cluster membership explained 67% of the total standardized variance in mOGTT-derived features (between-cluster sum of squares / total sum of squares, BSS/TSS = 0.675), with between-cluster variance far exceeding within-cluster variance (BCV = 235 vs WCV = 2), consistent with distinct glycemic phenotypes.

These more discernable, data-driven subgroupings also resulted in better alignment with 2IG-based classification. For instance, 100% of those in Group 3 (Severely Glucose Intolerant) had 2IG that exceeded the clinical 2-hour glucose threshold for PD (140 mg/dL) versus 83% under the A1c-based groupings.

These results indicate that clustering on mOGTT dynamics yields more physiologically coherent groups with more internal consistency, whereas A1c, the most commonly used marker of chronic average glycemia, groups together individuals with diverse glycemic response phenotypes. The high heterogeneity within the A1c-defined groups was most pronounced in T2D, supporting the interpretation that diverse glycemic response phenotypes are better captured by mOGTT- rather than by A1c-based groupings. More detailed descriptions of each of the three data-driven subgroups follows.

### Normoglycemia-Dominant Cluster (Glucose Tolerant; N=68)

This cluster exhibited glycemic features largely within the normal range (Figure 5B). The mean FIG was 102.75 mg/dL, closely aligned with the diagnostic cutoff for normal fasting glycemia (<100 mg/dL), with a range of 72 to 147 mg/dL. The average 2IG value during the OGTT was 121.25 mg/dL (range: 83 - 176 mg/dL), below the impaired glucose tolerance (IGT) threshold of 140 mg/dL, and mean AUC was 15,474.12 mg·min/dL. This group represents individuals with preserved glucose tolerance and low burden of metabolic abnormalities. Most of the normoglycemic participants (97%, N=34) and 24 PD and 10 T2D individuals fell in this category (Figure 5B), pointing to the presence of people with low-risk subphenotypes in these two A1c groups. The A1c values for individuals in this cluster ranged from 4.7 to 7.3, with a mean A1c of 5.7. (Figure 5C)

### Intermediate Dysglycemia Cluster (Glucose Intolerant; N=36)

This cluster demonstrated mOGTT values consistent with PD or early T2D. The mean FIG was 124.86 mg/dL (range: 94 - 179 mg/dL), approaching the T2D diagnostic cutoff of 126 mg/dL. 2IG averaged 190.94 mg/dL (range: 125 - 275 mg/dL), consistent with impaired glucose tolerance (IGT). AUC was elevated at 22,538.74 mg·min/dL on average, suggesting sustained post-load hyperglycemia. The relatively wide standard deviations observed (FIG: 17.54, 2IG: 37.06) point to clinical heterogeneity, underscoring the value of this cluster in capturing transitional glycemic states. A1c values in this group ranged from 5.5 to 8.3. Notably, 13 individuals with A1c-defined T2D and one with normoglycemia (with a relatively high A1c of 5.5) along with 22 PD individuals were categorized in this cluster, pointing to potentially preserved glucose load regulation in a subset of people who have A1c-defined T2D.

Relative to the A1c-defined PD group (n=46), the unsupervised Glucose Intolerant (Intermediate Dysglycemia) cluster (n=36) grouped together a markedly larger share of individuals exceeding OGTT-based dysglycemia thresholds. Specifically, FIG ≥100 mg/dL occurred in 94.4% of cluster members vs 73.9% in the A1c-defined PD group, and 2-hr glucose ≥140 mg/dL was present in 94.4% vs 56.5%, respectively. Exceedance of diabetes thresholds was also directionally higher in this cluster - FIG ≥126 mg/dL (50% vs 19.6%) and 2IG ≥200 mg/dL (41.67% vs 10.9%). This demonstrates multiple cases where A1c alone would misclassify early dysglycemia, but the data-driven cluster assignment would uncover “masked dysglycemia”, flagging high risk patients who would benefit from closer monitoring or earlier intervention.

### Severe Dysglycemia Cluster (Severe Glucose Intolerance; N=12)

Participants in this cluster exhibited marked glycemic impairment. The average FIG was 164.58 mg/dL (range: 131 - 225 mg/dL), well above the diagnostic threshold for T2D. 2IG values were profoundly elevated, averaging 282.92 mg/dL (range: 216 - 339 mg/dL), consistent with severe glucose intolerance. Relatedly, AUC was highest in this group at 29,978.38 mg·min/dL, indicating sustained and exaggerated postprandial hyperglycemia. This cluster represents a distinct phenotype of overt dysglycemia with high diagnostic certainty and metabolic burden and comprised only people from the A1c-defined T2D groups (n=12) and A1c between 6.5 and 8.7.

This cluster concentrated individuals with more extreme OGTT abnormalities than the A1c-T2D group. Every cluster member exceeded the PD threshold for 2-hr glucose (100% for this cluster vs 80% for A1c-T2D group) and everyone exceeded the diabetes threshold for 2-hr glucose (100% for this cluster vs 82.86% for A1c-T2D group). This indicates a markedly higher burden of 2-hr hyperglycemia in this cluster. PD-level elevation of FIG was near-saturated in both groups (100% for this cluster vs 97.1% for A1c-T2D group); T2D-level FIG was more frequent in this cluster (100.0% for this cluster vs 68.57% for A1c-T2D group)

The “Severe Glucose Intolerance” cluster captures a physiologically higher-risk phenotype than A1c-defined T2D, particularly for post-challenge hyperglycemia, a dimension tightly linked to cardiometabolic risk. While fasting abnormalities are common in both groups, the cluster substantially enriches for individuals crossing 2-hr PD and T2D thresholds, suggesting that unsupervised grouping on OGTT features reduces heterogeneity within the A1c-T2D category and may better flag patients who warrant intensified monitoring or earlier intervention. Replication in larger cohorts and prospective outcome links will be important to confirm clinical utility.

In all, the unsupervised clustering method offers a data-driven stratification approach, which we demonstrate maps onto traditional diagnostic categories while capturing within-group variability. The clear separation of clusters based on both fasting and post-load glucose metrics suggests that unsupervised learning using clinical thresholds for mOGTT metrics can effectively identify clinically meaningful subtypes.

## Discussion

This study demonstrates the feasibility, interpretation, and clinical relevance of CGM-based oral glucose tolerance tests (mobile OGTTs [mOGTT]) that can be performed anywhere, particularly, outside of clinical settings. We present the first large-scale, real-world evaluation of CGM-based OGTT (mOGTT) in individuals across the full glycemic spectrum, demonstrating expected groupwise dynamics and revealing new possibilities for stratifying metabolic risk. By evaluating high-resolution glucose challenge responses in a demographically diverse population (25.9% Black, 19.8% Hispanic), we show how mOGTT metrics can not only recapitulate known glycemic response dynamics but also reveal nuanced, subclinical glycemic subphenotypes that are not apparent from conventional diagnostics like A1c.

Importantly, we calculate data-driven diagnostic thresholds for CGM-based IFG and IGT that diverge from the default plasma-based criteria (higher for IFG and lower for IGT), differences which likely reflect the physiological time lag between interstitial and plasma glucose measurements during glycemic excursions^56^ and are consistent with literature showing that interstitial glucose levels are higher and dynamics are slower than plasma glucose during OGTT.^57^ Such differences between interstitial and plasma glucose underscore the need for CGM-specific diagnostic thresholds to enable effective clinical translation of mOGTTs.

Given that only blood-based thresholds exist to date, we applied those values to the CGM data during the mOGTT and saw substantial discordance between mOGTT and A1c-based diagnostic classifications, particularly in individuals with normoglycemia and PD. For example, 46% of A1c-defined normoglycemic participants met the FIG-based criteria for IFG. Conversely, a subset of participants with A1c-defined T2D demonstrated mOGTT profiles more consistent with PD or even normoglycemia, potentially indicative of glycemic improvement from recent lifestyle changes. These findings align with prior literature on diagnostic inconsistencies between A1c, fasting plasma glucose, and OGTT ^28,58^ and highlight the need for multimodal glycemic assessments to guide diagnosis and care.

Distinct glycemic pathophysiologies suggest that different lifestyle strategies may be more effective. IFG is characterized by early β-cell secretory failure on a background of hepatic insulin resistance: fasting glucose is elevated because hepatic glucose production is insufficiently restrained by insulin. By contrast, isolated IGT primarily reflects peripheral (skeletal muscle) insulin resistance with relatively preserved fasting glucose secretion but inadequate postprandial glucose disposal.^59^ Therapeutically, these distinctions suggest emphasis on preserving/augmenting β-cell function and suppressing hepatic glucose output in IFG (for example through moderate weight loss, caloric restriction and dietary patterns low in added sugars and refined starches)^60^ versus improving peripheral insulin sensitivity and postprandial control in IGT (e.g. prioritizing postprandial activity and structured exercise that increases skeletal muscle mass and oxidative capacity such as combined aerobic and resistance or interval training).^61–63^ Individuals exhibiting both IFG and IGT face the highest short-term risk. Concurrent fasting secretory defects and postprandial resistance together indicate a faster progression to T2D.^37^

The scalability of metabolic subphenotyping relies critically on the reproducibility of self-administered OGTTs outside of controlled clinical environments, as recently demonstrated by Metwally et al. (2024).^19^ Analysis of CGM traces from duplicate at-home sessions in the external cohort’s original analysis revealed high intra-individual consistency, with median Pearson correlation coefficient between two home-based glucose time series at r=0.86 (pvalue < 0.001).^19^ This robust temporal concordance suggests that the high variability often presumed in unsupervised settings can be effectively mitigated through protocol standardization. Specifically, strict adherence to detailed pre-test instructions (e.g., controlling preceding diet and physical activity) minimized intra-patient variability: the coefficient of variation (CV) for the home OGTT-derived CGM was just 11%^19^, comparing favorably to the published 16.7% CV for 2-hour plasma glucose during traditional OGTT.^64^ This confirmed reliability validates CGM-based OGTT performed outside of the clinic as a precise, high-throughput alternative to in-clinic OGTT for fine-grained metabolic classification.

Our findings support mOGTT as a viable and scalable tool for early dysglycemia detection in real-world settings. This aligns with the broader shift toward at-home care, remote monitoring, and precision prevention. Notably, we demonstrate that short-term glucose response metrics, particularly AUC and 1-hour glucose measured during the mOGTT, strongly correlate with longer-term CGM metrics such as GMI and time-in-range, especially in T2D individuals. These relationships suggest that mOGTT can serve as an indicator of both short- and longer-term glycemic control.

To address diagnostic inconsistencies and better capture the continuum of glycemic dysregulation, we developed the composite GCRI metric from the mOGTT that provides a practical, interpretable tool for real-world risk assessment and personalized intervention. Our findings support the GCRI as a marker of dysglycemia severity based on standard diagnostic metrics and thresholds. Further studies are needed to establish the GCRI as a clinically-useful biomarker and translate its diagnostic potential into practice.

An individual’s GCRI transforms CGM data collected during an mOGTT into actionable insights, not only by generating a single value that can be compared against normative thresholds and/or population spectra, but also by enabling dissection of the underlying pathophysiologies contributing to a person’s glycemic dysfunction. The GCRI’s constituent features provide a transparent fingerprint of underlying dysregulation, enabling clinicians to determine which physiologic domains are driving risk (e.g., IFG) and to tailor counseling or follow-up care accordingly. The GCRI showed superior performance to any single OGTT metric for diagnostic tasks, displayed stable generalization in an external cohort (N=29), and, importantly, detected masked dysglycemia that would likely have been missed by traditional clinical testing.

Leveraging unsupervised machine learning to “learn” subgroups of people with similar glycemic health characteristics revealed distinct glycemic and metabolic phenotypes that are independent of A1c. Two participants with very different physiological profiles (e.g., isolated fasting impairment vs. isolated post-load impairment) may have similar GCRI values, which motivated this unsupervised analysis. The clustering method identifies groups without forcing the use of traditional diagnostic cutoffs, improving granularity over A1c-based categories that are known to miss more subtle dysglycemia. This may aid in early detection, more personalized risk stratification, and titrating of interventions beyond conventional “one-size fits all” approaches based on diagnostic cutoffs. Conceptually, the GCRI provides a continuous, interpretable risk spectrum for individuals, while clustering of the underlying normalized differences between mOGTT metrics and their respective clinical thresholds reveals discrete subphenotypes with greater separability, offering practical implications for screening, triage, and personalized follow-up beyond A1c alone.

Limitations in this study include the use of A1c as the comparator to mOGTT which does not always serve as the best indicator of contemporary glycemic health. Further, this work used self–reported ingestion times for the mOGTT, which might not be completely accurate. However, we followed up with the participants shortly after the mOGTT to confirm the timings, which increases our confidence in correct timing. Future work will focus on external validation, calibration to outcomes, assessment across diverse populations, and refinement to better resolve mechanistically distinct subphenotypes. This can help individuals monitor their dysglycemia spectrum continuously, at-home conveniently. In the context of consumer-grade CGM availability – e.g., Dexcom Stelo and Abbott Libre Lingo^65^– mOGTTs present an actionable method for personal health tracking without requiring clinic visits or fingerstick measurements.

Looking ahead, the mOGTT lends itself to scalable, equitable implementation. A lightweight mobile interface paired with a CGM-enabled ecosystem could guide a standardized mOGTT at home (with timing prompts, start/stop capture), compute the GCRI in real time, and surface interpretable insights, comparisons to normative values, and give goal-oriented recommendations aligned to clinical guidance. Secure sharing with care teams would support longitudinal tracking, early detection of deterioration or improvement, and timely, targeted interventions. By lowering barriers to assessment by enabling fully remote assessment and by turning complex CGM data into a single interpretable index, this approach has the potential to improve access, personalize prevention, and support long-term metabolic and cardiovascular monitoring. Future studies should aim to validate these findings longitudinally and in diverse clinical settings, explore associations with cardiometabolic outcomes, and assess responsiveness to lifestyle and pharmacologic interventions such as GLP-1 receptor agonists. Our validation of the GCRI in the external cohort provides a roadmap for implementing GCRI in other external datasets or even as part of regular at-home patient monitoring.

As at-home monitoring technologies and AI-based digital health platforms become more accessible, mOGTT holds promise to form the foundation of next-generation diabetes risk assessment and intervention methods.

## Methods

### Study Population

Patients meeting eligibility criteria were recruited and enrolled from the Duke University Health System (DUHS) into the DiabetesWatch study (Pro00112384). We collected OGTT and smartwatch data from 116 individuals in the DiabetesWatch study (referred to as the DiabetesWatch population) in free-living settings using a CGM to measure glucose values. Eligibility criteria included being age 18 and above, with one A1c value in the past 6 months, and owning a mobile device that could connect to the study devices. Exclusion criteria included taking Metformin or other antidiabetic medications for the treatment of type 2 diabetes, currently on beta blockers, history of cardiovascular disease as evidenced by ASCVD (atherosclerotic cardiovascular disease), and patients with heart failure, history of cancer, COPD or chronic kidney disease (CKD), currently involved in a clinical trial that involves treatment, atrial fibrillation, or pregnancy. Of these participants, 35 were in the T2D group, 35 in the normoglycemic group, and 46 in the PD group. Participants were between the ages of 18 and 74 and were not on any anti-diabetic medications or insulin. A1c values ranged from 4.7 to 8.7.

### Devices and data collection protocol

For the DiabetesWatch study, data were collected over 14 days from a Freestyle Libre 3 CGM device, a Fitbit Sense 2 smartwatch, and an Empatica EmbracePlus smartwatch. PTS Diagnostics A1c Now kits were used for A1c determination at the end of participation (which was used for diagnosing norm, PD, and T2D). 590173 glucose readings were collected overall from the CGMs, which employ subcutaneous sensors to gauge interstitial fluid glucose levels at 5-minute intervals. Participants logged their diet and OGTT times using the Cronometer diet app with details on the time, calories, and composition of all food items consumed during this period. The first 24 hours of CGM data were not used for analysis due to higher errors reported during this time.

To evaluate diagnostic concordance, we computed class-wise sensitivity and specificity by comparing A1c-defined glycemic classification categories (norm, PD, T2D), which we treat in this analysis, against classifications derived from clinical FPG-based thresholds of FIG

To understand the feasibility and validity of performing an “at-home Oral Glucose Tolerance Test (OGTT)” using the CGM as a tool to capture measurements during the test, participants ingested 2 Boost beverages with total 74g carb load (480 cal) ^66^ in their homes after fasting (not synchronized at a specific morning hour) and following written guidance (Appendix 1).

### Outcomes

The primary objective was to define clinically relevant glycemic values across diagnostic groups spanning the dysglycemia spectrum (excluding individuals on medication or insulin) in free-living, at-home settings, and to develop a GCRI metric grounded in established clinical thresholds. Secondary objectives included examining the relationship between OGTT responses and established CGM metrics, understanding discrepancies in glycemic classification based on A1c versus OGTT-derived diagnostic metrics, and uncovering subphenotypes based on OGTT data.

### Data analysis

When levels are stable, interstitial glucose acts as a reasonable estimate of blood glucose.^35^ To aid clinically-relevant interpretation of CGM data, we explored applying clinical thresholds of FPG to FIG as a proxy for identifying Impaired Fasting Glucose (IFG) using CGM data. IFG is hallmarked by FPG ≥ 100 mg/dL, indicative of PD, or FPG ≥ 126 mg/dL, indicative of T2D.^1^ Thus, PD-level IFG is defined here as FIG between 100-126 mg/dL (represented by points falling between the yellow dashed and red dotted lines in Figure 2A and 2C), and T2D-level IFG is defined here as FIG >126 mg/dL (represented by points greater than the red dotted line in Figure 2D and 2F).

### OGTT Metrics Calculation

All data analysis was carried out in Python (version 3.12.4). OGTT features were calculated using the time of OTT start recorded in Cronometer and by the clinical research coordinator. OGTT features included standard time-point glucose values (e.g., 30-, 60-, and 120-minute glucose and fasting interstitial glucose (FIG)), derived indices such as peak glucose, area under the curve (AUC) (calculated using trapezoidal integration), incremental area under the curve (iAUC), and time to recovery (defined as the time to return to FIG). Standard time-point glucose values were determined by the glucose value recorded at the closest time within a 5-minute period to the target time. The mean, median, standard deviation, and min-max ranges of 6 metrics were calculated at the population level and each of the diagnostic group levels and reported in Table 1. These 6 metrics were chosen since they are routinely used in clinics for dysglycemia assessment (FPG, 30-minute, 1-hour, 2-hour glucose post OGTT), and AUC and iAUC are increasingly being explored for risk stratification. For participants who did not return to their FIG value in the 120-min observation window, we imputed a time-to-return of 120 minutes and then calculated mean time to baseline per group. The clinically and research-established thresholds for glucose tolerance were used to determine how many individuals in each category lay outside the threshold based on their OGTT metric. Pearson correlation of each metric with GMI was also reported, where GMI was calculated using the iglu_py package (version 1.1.1).

### Data-driven cutoffs for CGM-based IFG classification

To quantitatively assess the alignment between fasting glucose (FIG) and A1c-based diagnostic categories, we conducted a threshold optimization analysis using a grid search strategy. Each individual in the dataset was first categorized into one of three glycemic states - norm, PD, or T2D - based on their A1c values, using established clinical cutoffs of <5.7%, 5.7–6.4%, and ≥6.5%, respectively. We then evaluated a range of candidate FIG thresholds to determine the pair of cut points that most accurately recapitulated this classification.

The optimization procedure systematically varied the lower FIG threshold (used to distinguish normal glucose regulation from PD) from 85 to 110 mg/dL, and the upper threshold (used to separate PD from T2D) from 115 to 140 mg/dL, both in increments of 0.5 mg/dL. For each threshold pair, we reassigned glycemic categories based solely on FIG values: individuals with values below the lower threshold were labeled as normoglycemic, those between thresholds as having PD, and those above the upper threshold as diabetic. These FIG-derived classifications were then compared against the A1c-based labels, and the classification accuracy, defined as the proportion of individuals whose FIG-based label matched their A1c-based diagnosis, was computed for each threshold combination. The classification accuracy for each threshold combination was computed as the mean of these binary matches, effectively representing the proportion of individuals for whom the FIG-derived label was identical to the A1c-based label. This method treats each subject equally and yields a robust measure of overall alignment across the cohort. We used multi-class confusion matrices to quantify true and false positive rates for each classification method, treating A1c as a ground truth to calculate sensitivities and specificities for norm, PD, or T2D.

To extend this approach to postprandial glucose dynamics, we applied a parallel threshold optimization analysis using 2IG values. Using the same A1c-based diagnostic groups (norm: <5.7%, PD: 5.7–6.4%, T2D: ≥6.5%) as the reference standard, we systematically searched for the pair of 2IG thresholds that best reproduced these categories. The lower 2IG cutoff, intended to distinguish normoglycemia from prediabetes, was varied from 120 to 160 mg/dL, while the upper cutoff separating PD from T2D ranged from 180 to 220 mg/dL, again in 0.5 mg/dL increments. For each threshold pair, individuals were assigned a glucose-derived diagnostic label based solely on their 2IG values. Accuracy was computed as the proportion of cases where these 2IG-derived labels matched the A1c-based classification. This method enabled a direct comparison of the discriminatory performance of fasting versus post-load glucose measurements relative to A1c-defined glycemic states. Thresholds used to define dysglycemia for mOGTT peak glucose and peak onset timing were set at the 10th percentile of the PD distribution, estimated using empirical quantiles.

### GCRI Using mOGTT Features and Clinical Thresholds

To quantify glycemic dysregulation across multiple metabolic dimensions, we developed a GCRI based on deviations from clinically established thresholds. We selected six physiologically relevant and clinically established features derived from the oral glucose tolerance test (OGTT): fasting plasma glucose (here, FIG), 30-minute glucose, 1-hour glucose, 2-hour glucose (here, 2IG), area under the glucose curve, and glucose value difference between 2 hours and start of OGTT.^1,17^

Each feature has units of mg/dL and is calculated as the difference between the measurement taken as a part of the mOGTT test and the clinical or prior–research–based thresholds (for AUC) for glucose intolerance and T2D:

1. FIG −100
2. 30-min glucose delta from 165.6 mg/dL
3. 1-hour glucose delta from 155 mg/dL
4. 2-hour glucose delta from >=140 mg/dL
5. AUC (calculated using trapezoidal rule): 18,300 mg·min/dL
6. End-to-start glucose value difference: 0 mg/dL

Additionally, we measured different types of dysglycemia from mOGTT (based on mOGTT metrics such as FIG, 2IG etc.) and present an accompanying heatmap for the mOGTT score. Dysglycemia was defined as those metrics exceeding respective PD thresholds. For AUC and iAUC, we also report dysglycemia (high AUC and iAUC) where these metrics exceeded the respective mean cohort values. Reactive hypoglycemia and rapid drop in glucose after OGTT were also assessed based on whether the glucose value plunged below FIG in the first 15 minutes during OGTT and whether the glucose value plunged below FIG at least twice during mOGTT, respectively. To quantify an individual’s overall deviation from normative glycemic responses, we developed GCRI, incorporating standard OGTT-derived features and their deviations from diagnostic glucose intolerance and diabetes thresholds.

For each metric, a clinically relevant reference threshold described above was used based on ADA and IDF criteria.^9^

To compute the GCRI, we first calculated standard deviation for each feature f: 𝜎_𝑓_ = 𝑆𝑡𝑑 (𝑋_𝑓_);(Ddof = 1)

The standardized deviation from threshold per participant i for each feature f was then calculated as: 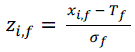

The raw GCRI mean across features was calculate (with F the set of the 6 features above) as:

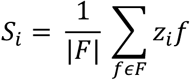

The score was the scaled between 0 and 1. Let S_min_ = min_i_ S_i_ and S_max_ = max_i_ S_i_, the GCRI was calculate as:

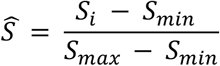

Classification performance of the GCRI was evaluated as the proportion of individuals whose predicted diagnostic category (norm, PD, or T2D) matched their A1c-based diagnostic category.

Participant values were standardized as z-scores based on population-level standard deviations and their distance from the thresholds. These standardized deviations were then averaged across all features to generate a GCRI capturing the cumulative dysglycemia burden. To enhance interpretability, the final score was linearly scaled to a bounded range [0, 1], where 00 indicates healthy physiology and 1 reflects the greatest relative glycemic dysregulation. This unified measure enables integrated risk stratification across the prediabetes-to-diabetes continuum. To establish clinically meaningful cutoffs, we performed a data-driven threshold optimization using A1c-based diagnosis as the reference standard (normoglycemic <5.7%, PD 5.7–6.4%, T2D ≥6.5%). Candidate thresholds for separating normoglycemic vs. PD and PD vs. T2D were scanned across the empirical distribution of GCRIs, and the pair maximizing overall accuracy was selected. Individuals were then classified into norm, PD, or T2D categories based on their position relative to the optimal thresholds.

We externally validated the GCRI on an independent OGTT dataset (details can be found in the original manuscript cited)^19^ by extracting the required features - fasting (0 min), 60-min, 120-min glucose via the nearest measurement within ±5 min, the 0-120 min AUC computed by trapezoidal rule using only observed points with measured endpoints within ±5 min, and the 2h–baseline difference. These features were projected onto the DiabetesWatch cohort’s scaling (feature-wise deviation from the pre-specified thresholds divided by the original cohort SDs), averaged to a GCRI, and linearly rescaled to the original cohort’s range. DI was calculated for this cohort using insulin secretion rate using the C-peptide deconvolution method on C-peptide concentrations measured during OGTT tests at 0, 15 and 30 min, as explained in the original research paper.^19^

### Cluster Identification

To uncover data-driven glycemic phenotypes, we applied unsupervised clustering to individuals based on the above-described deviations from clinically meaningful glucose thresholds across key time points in the oral glucose tolerance test (OGTT). To recover richer structure while retaining clinical interpretability, we represented each participant by their difference-from-threshold profile across the full mOGTT panel and performed unsupervised clustering on these multi-feature vectors (with a leave-one-metric-out setup for unbiased evaluation of each withheld metric). In practice (e.g., K-means), clusters minimize within-cluster dispersion and maximize between-cluster separation, yielding groupings that reflect how glycemia is dysregulated (fasting-dominant, post-load-dominant, mixed, etc.). This approach thus uses the same clinically anchored scaling as the GCRI but keeps direction and magnitude across all metrics. K-means clustering was applied to the normalized deviation matrix. To determine the optimal number of clusters (k), we computed the within-cluster sum of squares (inertia) for values of k ranging from 1 to 9. The elbow method^67^ was used to visually assess the point of diminishing returns, and the optimal number of clusters (k = 3) was selected based on the inflection point in the inertia curve. The cluster labels were used to stratify individuals into glycemic subtypes.

Clustering analyses were performed on the same six mOGTT-derived variables: FIG, 30-min glucose, 1-hr glucose, 2IG, AUC, and the end-to-start change in glucose. Similar to GCRI calculation, the distance of each variable was calculated from prespecified thresholds (FIG 100 mg/dL; 30-min 165.6 mg/dL; 1-hr 155 mg/dL; 2IG 140 mg/dL; AUC 305×60 mg·min/dL; end–start glucose difference 0 mg/dL) and then standardized by the cohort standard deviation for that metric.

We transformed each mOGTT feature into a threshold-anchored standardized deviation (X_i,f_ – T_f_ / σ_f_. Using these vectors, we fit K-means (elbow-guided k) on: (i) the full feature set and (ii) a series of leave-one-metric-out variants (e.g., “drop FPG,” “drop 1-hr,” …). For each variant, we then examined the withheld metric’s empirical distribution across the learned clusters (KDEs/histograms), overlaid clinical thresholds, and computed the percentage of subjects from each group exceeding PD thresholds per cluster.

Unsupervised clustering uses the same per-feature deviations that feed into the GCRI, but retain the full vector of mOGTT metric distances for each participant:

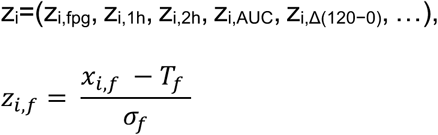

where i is the: index for participants and f is the index for metrics (6 total), f∈F.

Grouping participants in this 6-dimensional space preserves the pattern and magnitude across features, rather than collapsing them to a single average.

Unsupervised clustering based on the differences of the mOGTT metrics from respective clinical thresholds as mentioned in mOGTT methods above (FIG: 100 mg/dL, 30-min glucose: 165.6 mg/dL, 1-hour glucose: 155 mg/dL, 2-hour glucose: 140 mg/dL, AUC: 18,300 mg·min/dL, End-to-start glucose value difference: 0 mg/dL) revealed three distinct subgroups using the elbow method, each with unique metabolic profiles that align with established diagnostic categories for glucose dysregulation.

We quantified the homogeneity within clusters using a variance-based purity metric computed on mOGTT features. We compared two grouping schemes: (i) GlycemiaCluster, a data-driven three-cluster solution obtained by k-means on the threshold-centered, standardized OGTT feature space; and (ii) A1c based diagnosis, the A1c-defined diagnostic categories (norm, PD, Diab) as present in the dataset. For each scheme, we computed variance-based purity separately for every group and every feature. Purity was defined as the reduction in within-group variance relative to the overall cohort variance for that feature, so that higher values indicate tighter, more homogeneous groups on the corresponding OGTT dimension. When within-group variance exceeded the cohort variance, the resulting value was truncated at zero for reporting consistency.

To quantify group homogeneity, we used a variance-based purity metric computed per group g and feature f as

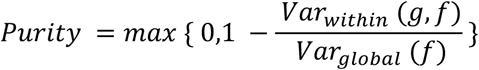

Here, x_i,f_ denotes subject i’s value for feature f.

The global variance

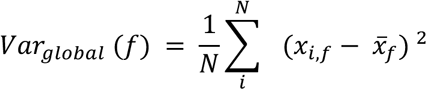

measures dispersion across the entire cohort, and the within-group variance

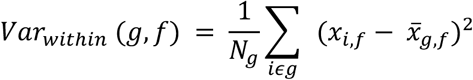

measures dispersion among members of group g.

When within-group variance exceeded the cohort variance, the resulting value was truncated at zero for reporting consistency. A higher purity value indicates that the group’s spread on that feature is a smaller fraction of the cohort spread (i.e., tighter, more homogeneous), approaching 1 when within-group variance is much smaller than global variance and approaching 0 when it is comparable; ratios exceeding 1 are clipped to 0 for reporting. For each grouping scheme, we formed per-feature, cluster-size-weighted purities by averaging group purities with weights n_g_ / N, then averaged these across features to obtain a single cluster-level purity, yielding the reported values (e.g., 0.662 for unsupervised clusters vs 0.381 for A1c group).

To summarize each scheme at the feature level, we formed a cluster-size-weighted purity, i.e., a weighted average of group purities using group sample sizes as weights, yielding one purity value per feature for each scheme. A single scheme-level purity was then obtained by averaging these per-feature values across the six OGTT metrics. In addition to the scheme-level summary, we report group-level purities to show how homogeneity varies across clusters or diagnoses, and per-feature, cluster-weighted purities to identify which OGTT dimensions contribute most to differences between schemes. All computations were implemented in Python (pandas (version 2.2.2), NumPy (version 1.26.4), scikit-learn (version 1.5.1), using population variance estimates and a fixed random seed for reproducibility of the clustering step.

For each clustering solution, we also computed the overall mean of the feature matrix and the mean feature vector (centroid) within each cluster. Using these, we decomposed the total variability of the standardized features into within-cluster and between-cluster sums of squares. From this decomposition, we derived pooled within-cluster variance (WCV), pooled between-cluster variance (BCV), and the proportion of total variance explained by the clusters (between-cluster sum of squares divided by total sum of squares). We also summarized the variance ratio using the Calinski–Harabasz index, which compares between-cluster dispersion to within-cluster dispersion while accounting for the number of clusters and sample size.

As a distance-based, point-level measure of cohesion and separation, we computed the Silhouette coefficient for each participant based on Euclidean distances in the standardized feature space and reported the mean Silhouette score across all participants for the selected 3-cluster solution.

### Leave-one-metric-out (LOMO) clustering: rationale and design

To rigorously evaluate how each mOGTT metric behaves across pathophysiologic groups, without baking that metric into the group definitions, we used a leave-one-metric-out (LOMO) clustering strategy. The key idea is to avoid circularity: if clusters are formed using a given metric, then inspecting that same metric’s distribution across those clusters can be tautologically favorable. LOMO prevents this by withholding the metric under evaluation when deriving the clusters. mOGTT captures multiple, partially redundant dimensions of dysglycemia (fasting level and recovery, early/peak/post-challenge responses, total/iAUC).

We constructed threshold-anchored feature deviations for the full mOGTT panel, computing (metric value − threshold for that metric) / σ for that metric, for each metric. Then, for each metric M in turn (FPG, 30-min, 1-hr, 2-hr glucose, AUC, and iAUC), we removed M and fit a K-means model to the remaining features. To ensure interpretability across these leave-one-metric-out variants, cluster labels were aligned yielding consistent names: Glucose Tolerant, Glucose Intolerant, and Severe Glucose Intolerant. We evaluated the withheld metric M across the resulting clusters using distributional summaries (violin/KDE plots) and clinical exceedance analyses (proportions at or above prediabetes and diabetes thresholds). This procedure was repeated for every metric; for iAUC we additionally reported behavior under the “ALL-features” model to provide composite context.

We transformed each OGTT feature into a threshold-anchored standardized deviation (X_i,f_ – T_f_ / σ_f_. Using these vectors, we fit K-means (elbow-guided k) on: (i) the full feature set and (ii) a series of leave-one-metric-out variants (e.g., “drop FPG,” “drop 1-hr,” …). For each variant, we then examined the withheld metric’s empirical distribution across the learned clusters (KDEs/histograms), overlaid clinical thresholds, and computed the percentage of subjects from each group exceeding PD thresholds per cluster.

By withholding (M), any separation observed for (M) is earned by correlated physiology captured in the *other* features, not by construction providing Unbiased comparison. Using clinically researched thresholds to compute ≥PD and ≥T2D proportions ties cluster differences directly to decision-making rather than purely statistical contrasts. LOMO mimics prospective use, where a metric may be noisy or missing; clusters should remain informative when any single feature is unavailable.

For each violin plot panel, we visualize the metric’s distribution per cluster, annotate the percent ≥PD, and compare these to A1c-defined strata as a pragmatic benchmark. Concordant findings across LOMO variants indicate that clustering captures a multidimensional dysglycemia phenotype rather than a single-marker artifact.

To rigorously evaluate how each mOGTT metric behaves across pathophysiologic groups, without baking that metric into the group definitions, we used a leave-one-metric-out (LOMO) clustering strategy. The key idea is to avoid circularity: if clusters are formed using a given metric, then inspecting that same metric’s distribution across those clusters can be tautologically favorable. LOMO prevents this by withholding the metric under evaluation when deriving the clusters, preventing optimistic separation for any one marker. LOMO provides a built-in negative control that reduces information leakage, yields a more unbiased estimate of metric–cluster associations, and highlights which signals persist even when a metric is not allowed to drive the grouping.

## Supporting information

Supplementary Image 1, Supplementary Image 2, Appendix 1

## Data Availability

To enable others to validate this work and carry out similar research, the mOGTT study protocol including participant-facing instructions are available in the Supplementary Information. All data used for the analysis presented herein, specifically including the de-identified continuous glucose monitor data collected during the mOGTT, alongside relevant metadata like A1c and demographic information, will be made available on PhysioNet. To facilitate reproducibility, all code generated and used for this research, including detailed information about coding and compute environment, will be made available on https://github.com/Big-Ideas-Lab.

---

**Box 1.** Glossary of relevant terms and acronyms either already used clinically or introduced by this work.

| Term | Definition | Biological Range<sup>1</sup> |
| --- | --- | --- |
| mOGTT | Mobile OGTT: OGTT performed using CGM | N/A |
| GCRI | Glucose Challenge Response Index | N/A |
| FIG | Fasting Interstitial Glucose (analogous to Fasting Plasma Glucose in traditional OGTT); used to identify IFG | *Normoglycemia: 72 - 126<br>*PD: 84 - 179<br>*T2D: 95 - 225 |
| 30IG | Interstitial glucose value at 30-min post glucose load (equivalent to 30-min plasma glucose in traditional OGTT) | *Normoglycemia: 101 - 187<br>*PD: 107 - 232<br>*T2D: 133 - 284 |
| 1IG | Interstitial glucose value at 1-hour post glucose load (equivalent to 1-hour plasma glucose in traditional OGTT) | *Normoglycemia: 83 - 250<br>*PD: 85 - 250<br>*T2D: 112 - 328 |
| 2IG | Interstitial glucose value at 2-hour post glucose load (equivalent to 2-hour plasma glucose in traditional OGTT); used to identify IGT | *Normoglycemia: 88 - 165<br>*PD: 83 - 258<br>*T2D: 106 - 339 |
| AUC | Area Under Curve: Total area under the 2-hour mOGTT glucose curve | *Normoglycemia: 10547.5 - 23425.0<br>*PD: 12115.0 - 25580.0<br>*T2D: 16275.5 - 34292.5 |
| iAUC | Incremental area under curve: Incremental area under the 2-hour mOGTT glucose curve | *Normoglycemia: 1350 - 11465<br>*PD: 595 - 12007.5<br>*T2D: 1754 - 14517.5 |
| NGT | Normal Glucose Tolerance: Normal fasting glucose levels and normal glucose tolerance | FIG~FPG < 100 and 2IG~2-hour glucose < 140 |
| I-IFG | Isolated Impaired Fasting Glucose: Impaired fasting glucose levels in the absence of IGT | I-IFG: $100 \leq \text{FIG} \sim \text{FPG} < 126$ and 2IG~2-hour glucose < 140 |
| I-IGT | Isolated Impaired Glucose Tolerance: Impaired glucose tolerance with normal fasting glucose levels | FIG~FPG < 100 and $140 \leq 2\text{IG}\sim 2\text{-hour glucose} < 200$ |
| I-IFG+T2D | I-IFG, including IFG indicative of T2D | FIG~FPG $\geq 100$ and $2\text{IG}\sim 2\text{-hour glucose} < 140$ |
| I-IGT + T2D | I-IGT, including IGT indicative of T2D | FIG~FPG < 100 and $140 \leq 2\text{IG}\sim 2\text{-hour glucose}$ |
| Combined IGT and IFG (including T2D level) | IFG, including IFG indicative of T2D, and IGT, including IGT indicative of T2D | FIG~FPG $\geq 100$ and $2\text{IG}\sim 2\text{-hour glucose} \geq 140$ |
| FIG indicative of PD | FIG levels that indicate PD | $100 \leq \text{FIG}\sim\text{FPG} < 126$ |
| FIG indicative of T2D | FIG levels that indicate T2D | FIG~FPG $\geq 126$ |
| Peak Glucose Dysglycemia | Peak Glucose at or above 200 mg/dl | Peak glucose $\geq 200$ |

\*Based only on DiabetesWatch mOGTT data

## Acknowledgements

We would like to thank Abha Singh, Wan Lan Liang, Kaileigh Moertl, and Stephanie Sullivan for their valuable efforts throughout the project. This work was supported by NIH/NIDDK R01 DK133531 award. We acknowledge the support of the Duke Clinical and translational science Institute.

## Author Contributions

K.S. designed the study, wrote the manuscript, performed computational analyses and developed visualizations. J.D., L.E., M.J.C., and A.A. provided guidance on data analysis and clinical interpretation. A.B. and K.S. collected mOGTT data from DiabetesWatch participants. P.N., K.S., and A.B. helped assess data missingness. J.D., B.B. and K.S. helped with study design. L.H., B.C., S.M.I., B.B., P.J.C., M.M.S., and W.K.W. assisted with data interpretation. A.A.M. and M.P.S. provided data used in the external dataset analysis and provided guidance on interpretation. K.S. and J.D. wrote the manuscript with input from all authors.

