## Supplementary Image 1, Supplementary Image 2, Appendix 1 for "Unmasking Hidden Dysglycemia: A Mobile OGTT Approach Using Continuous Glucose Monitors"

**A**

| | Pred Norm | Pred PD | Pred T2D |
|---|---|---|---|
| True Norm | 27 | 8 | 0 |
| True PD | 9 | 35 | 2 |
| True T2D | 0 | 13 | 22 |

**B**

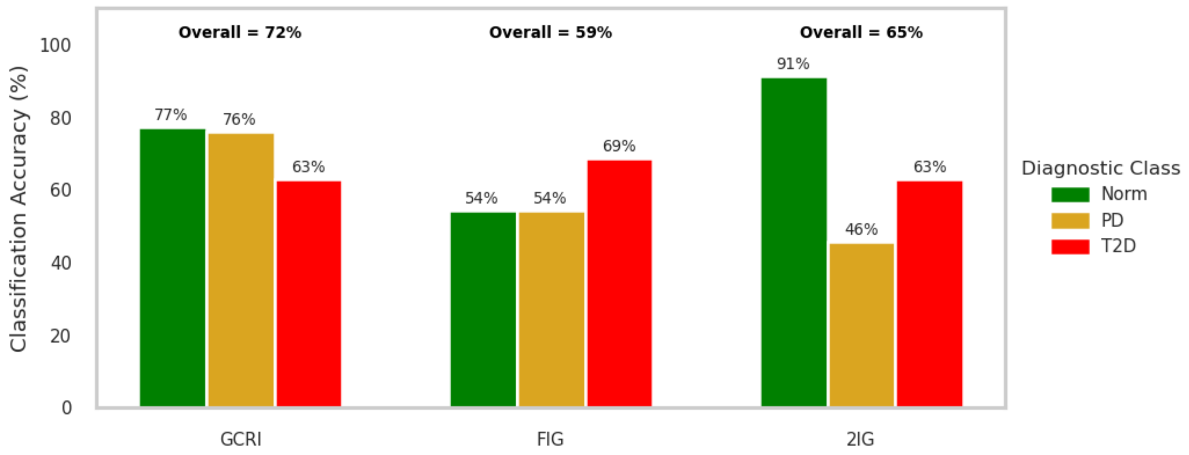


**Supplementary Figure 1. A. Confusion matrix for GCRI–based classification relative to A1c-defined diagnostic categories.** Each cell indicates the number of individuals assigned to a predicted category (columns) given their true category based on A1c (rows). Class-specific accuracy was highest for PD and Norm, while the majority of misclassifications involved individuals with T2D being assigned to the PD group.

**Supplementary Figure 1.B. Comparison of classification accuracy by method (GCRI, FIG, and 2IG).** Bars represent class-specific accuracy (proportion correctly classified within each true diagnostic category), with colors denoting the true class: green = Norm, yellow = PD, red = T2D. Overall accuracy for each method is annotated above its group. The GCRI achieved the highest overall accuracy and provided the most balanced performance across categories compared to FIG and 2IG thresholds.

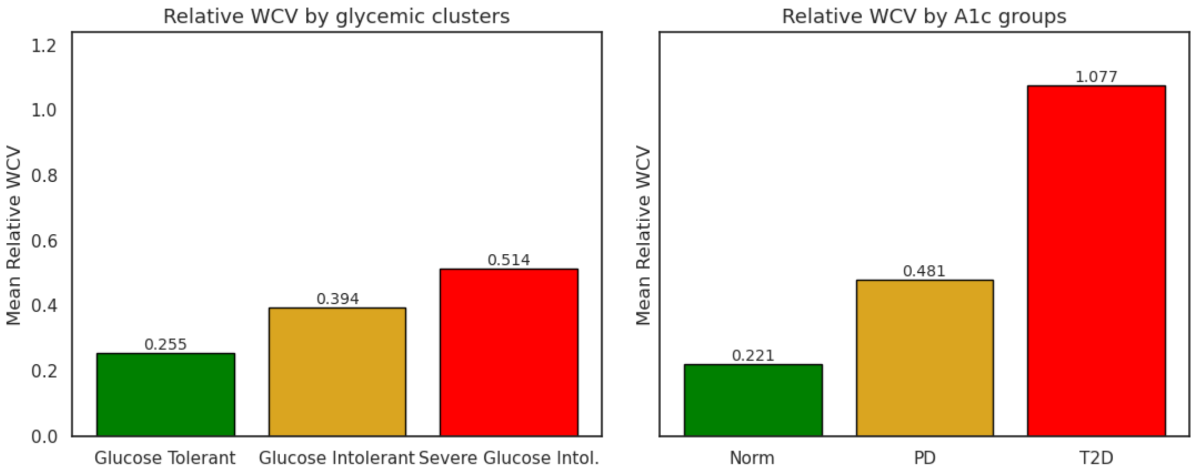


**Supplemental Figure 2. Mean relative within-cluster variance (WCV) for glycemia clusters and diagnostic groups.** Bars represent the mean relative WCV, calculated as the average ratio of within-group to global feature variance across all mOGTT-derived metrics. Lower values indicate tighter, more homogeneous groups.

**Appendix 1.**

### Oral Glucose Tolerance Test Instructions

#### (To be performed on xx date first thing in the morning)

1. Perform the test on an empty stomach in the morning, shortly after waking up. You must not consume anything 8 hours prior to the test. Do not consume anything else besides the Boost until the end of the standardized meal test.
2. You will need to be seated for 2 hours so choose a comfortable seating position and location to relax.
3. Note down the start time of the test after you sit down and right before you drink the beverage (Boosts). Note this down on the instruction manual in the space provided below. Please ensure you note the times accurately down to the minute. You will be asked to share the time with the research team at the end of the test.
4. Open both the boost bottles and drink the whole beverage from both bottles (one after the other) at a normal pace until the drinks are finished. Please consume the drinks from both bottles in total 5 minutes or less. The drink can be consumed at room temperature or cold according to your preference.
5. Note the end time after you finish drinking the Boosts in the space provided below. At this time, record the Boost drinks in your Cronometer app as well.
6. Sit still for two hours from the time you finish drinking the Boosts. You can choose to listen to music, read, play games, or watch TV or a movie during this time. However, please ensure you do not do something that causes too much anxiety, excitement, or anger as this can affect the data. At the end of your oral glucose tolerance test time, note down the time you get up in the space provided below.
7. You will receive a call from our research team shortly after. You will be asked to tell them the time when you started the test (right before you started drinking the Boost), the time you ended drinking the Boost, and the time you got up at the end of the 2 hours.
8. You can get up to visit the bathroom if you have to but make sure to walk slowly and sit down right after. If you get up, please note the times and frequency you got up for the bathroom, etc.

#### OGTT Time Log

| Event Name | Time (HH:MM) |
|---|---|
| What time did you start drinking Boost? | |
| What time did you finish drinking Boost? | |
| What time did you get up after the test? | |

*Please note below if you got up for any reason, for example, to use the restroom, and what time(s) you got up.*

*Please note below what you were doing during the OGTT. For example, worked, watched a movie, read a book, etc.*
